# Associations between plasma proteins and psychological wellbeing: evidence from over 20 years of the English Longitudinal Study of Ageing

**DOI:** 10.1101/2025.02.05.25321715

**Authors:** Jessica Gong, Shaun Scholes, Steven Cole, Paola Zaninotto, Andrew Steptoe

**Affiliations:** Department of Epidemiology and Public Health, University College London, London, UK; George Institute for Global Health, Imperial College London, London, UK; Department of Neurobiology, Care and Society, Karolinska Institutet, Stockholm, Sweden; Cousins Center for Psychoneuroimmunology, Semel Institute for Neuroscience and Human Behavior, and Department of Psychiatry & Biobehavioral Sciences, University of California, Los Angeles (UCLA) School of Medicine, Los Angeles, CA, USA; Department of Behavioural Science and Health, University College London, London, UK

## Abstract

A deeper understanding of the molecular processes involved in psychological wellbeing in older adults is essential for advancing knowledge of underlying biological mechanisms. Leveraging proteomics data from 3,262 older adults (mean age=63.5 years, 55% female) of the English Longitudinal Study of Ageing (ELSA), we investigated the cross-sectional and longitudinal associations (before and after protein measurement) between 276 proteins and eudaimonic wellbeing, hedonic wellbeing, life satisfaction, and depressive symptoms, over 20-year span. For positive wellbeing, two proteins (DEFB4A and ECE1) were longitudinally associated with subsequent eudaimonic wellbeing trajectory. We further identified higher concentrations of 7, 8, and 2 proteins were linked to subsequent lower eudaimonic wellbeing, hedonic wellbeing, and life satisfaction, respectively. Sex differences in XCL1 and SLAMF7 were observed, associated with lower eudaimonic and hedonic wellbeing in males. These findings link human psychological wellbeing to regulation of several biological pathways, particularly involving cytokine regulation, neurotrophic signaling, inflammatory and immune systems.

## INTRODUCTION

Psychological wellbeing has become a major issue in public policy, economics, and health over the past two decades.[1] There is substantial evidence that depression and other negative emotional states are associated with the development and prognosis of multiple health outcomes.[2] Positive psychological wellbeing appears to have a protective effect on health independently of negative emotions.[3] This has led to the search for biological pathways underpinning links between positive wellbeing and health outcomes.

Positive psychological wellbeing is a complex and multifaceted concept, and involves three related constructs.[1] First, evaluative wellbeing encompasses overall life satisfaction, whether on a general level or in specific aspects of one’s life. Second, hedonic or affective wellbeing relates to mood and emotional experiences of joy and happiness, and the philosophical conception of hedonism posits the pursuit of pleasure and the minimization of pain as the ultimate ends of human life.[4] Third, eudaimonic wellbeing extends beyond pleasure and pain, emphasizes the sense of purpose and meaning in life,[5] and the pursuit of self-determination.[6] Although eudaimonic wellbeing is often characterized as a more in-depth dimension of wellbeing,[7, 8] empirical evidence indicates that it is strongly correlated with hedonic wellbeing, and the two constructs appear to exert reciprocal influences on one another.[4] All three concepts have been found in epidemiological cohorts to be associated with longer survival, and the postponement of serious conditions such as coronary heart disease and disability at older ages.[9]

Studies of the biological correlates have tended to concentrate on a single dimension of psychological wellbeing, and to focus on processes previously identified in stress research as sensitive to emotional states; these include reduced levels of cortisol output, reduced cardiovascular responses to acute challenge, and lower concentrations of inflammatory markers.[3, 4] A gene expression analysis suggests that the molecular underpinnings of eudaimonic and hedonic wellbeing may be distinct: using the conserved transcriptional response to adversity (CTRA) as a molecular framework to investigate differential biological correlates, eudaimonic wellbeing was characterized by downregulation of the CTRA transcriptome, reflected in decreased expression of genes involved in proinflammatory signaling and increased expression of genes supporting antiviral and antibody-related responses. In contrast, higher levels of hedonic wellbeing were associated with relative upregulation of the CTRA profile, including increased transcription of proinflammatory cytokine genes.[8, 10] Although this literature provides valuable insight, most biomarker studies have been candidate-driven and have rarely extended to proteome-wide approaches. Existing proteomic investigations using plasma samples in mental health have primarily examined mood disorders and depression,[11, 12] with little attention to positive psychological wellbeing. Additionally, sex differences have been inconsistently reported,[13] and bidirectional associations may be present.

This study employed a data-driven approach utilizing high-throughput proteomics data to explore associations with trajectories of distinct dimensions of wellbeing, such as hedonic wellbeing, eudaimonic wellbeing, life satisfaction (or evaluative wellbeing) as well as depressive symptoms and depression. By embedding analyses in the English Longitudinal Study of Ageing (ELSA), we were able to not only assess the cross-sectional associations between protein measures and psychological wellbeing, but also to study the relationships with wellbeing trajectories longitudinally, recorded in the years prior to, and up to 15 years after the protein assays, spanning approximately 20 years. This approach allowed us to explore the relationships between protein biomarkers and psychological wellbeing before, during, and after the protein measurements.[14]

## RESULTS

The participant selection for the proteomics assays in ELSA is presented in Supplementary Figure 1. A full list of the 276 proteins assayed (Unitprot ID, gene and protein name) is included in Supplementary Table 1. At ELSA study wave 4 in 2008, the participants included in the current study had a mean age of 63.5 years [standard deviation (SD)=9.4], with 55% being female and 97% of White ethnicity (Table 1). 5.2%, 1.6%, 1.7%, 4.6%, 19.4%, and 2.0% had self-reported physician-diagnosis of cardiovascular disease (angina, heart attack, heart failure), stroke, cancer, diabetes, arthritis, and chronic lung disease at wave 4, respectively.

The wave 4 mean score for: 1) ***eudaimonic wellbeing*** (ranged 0-42, with higher score reflecting higher levels of wellbeing) measured using the 15 items of Control, Autonomy, Self-Realization and Pleasure (CASP) was 28.6 [SD=6.9]; for 2) ***hedonic wellbeing*** (ranged 0-12, with higher score reflecting higher wellbeing) measured using the pleasure subscale of CASP was 10.0 [SD=1.8]; for 3) ***life satisfaction (or evaluative wellbeing)*** (ranged 0-30, with higher score reflecting higher wellbeing) measured using the Diener Life Satisfaction scale (SWLS) was 20.4 [SD=6.2]; and 4) ***depressive symptoms*** (ranged 0-8, with higher score reflecting a higher number of depressive symptoms) measured using the Center for Epidemiological Studies-Depression scale (CESD) was 1.3 [SD=1.8], with 18.1% defined as having depression using the 3.0 cut-off point on CESD.

Correlation matrices showing the Spearman’s rank correlation coefficient (ρ) between wellbeing measures and depressive symptoms by study wave were included in Supplementary Figure 2. The strongest correlations were observed between eudaimonic and hedonic wellbeing across waves (ρ=0.48 to 0.70), indicating a positive relationship between these two measures of wellbeing. Additionally, moderate positive correlations were found between eudaimonic wellbeing and life satisfaction across waves (ρ=0.45 to 0.61).

### Analysis 1: Cross-sectional protein-wellbeing associations at wave 4

At Wave 4 in 2008, results from the cross-sectional linear regression models after minimal adjustments for age, sex, wealth quintile, and ethnicity are shown for eudaimonic wellbeing (Supplementary Figure 3A; Supplementary Table 2), hedonic wellbeing (Supplementary Figure 3B; Supplementary Table 3), life satisfaction (Supplementary Figure 3C; Supplementary Table 4) and depressive symptoms (Supplementary Figure 3D; Supplementary Table 5).

After further adjustment for the presence of six physician-diagnosed chronic diseases (cardiovascular disease, arthritis, cancer, diabetes, chronic lung disease, and stroke), and smoking status, the results revealed that higher eudaimonic wellbeing was associated with lower concentrations of 14 proteins (TNFRSF10A, KIM1 (also known as HAVCR1), EDA2R, ASGR1, TNFRSF11A, GFR.alpha.1 (also known as GFRA1), UNC5C, FGF.23, AMBP, Gal.9 (also known as LGALS9), EFNA4, MSR1, SCARB2, CD302; Figure 1A; Supplementary Table 6), and higher hedonic wellbeing was associated with lower concentrations of 14 proteins (TNFRSF10A, ASGR1, GFR.alpha.1, EDA2R, UNC5C, TNFRSF12A, TNFRSF11A, SCARB2, VSIG2, KIM1, EPHB6, AMBP, MSR1, SKR3 (also known as ACVRL1; Figure 1B; Supplementary Table 7) (all P_FDR_ < 0.05). No protein was associated with life satisfaction (Figure 1C; Supplementary Table 8), and we did not find any proteins that showed a positive association (increased abundance) with any dimension of wellbeing. No protein was associated with the level of depressive symptoms (Figure 1D; Supplementary Table 9), nor with depression (Supplementary Figure 4A; Supplementary Table 10).

**Figure 1.**
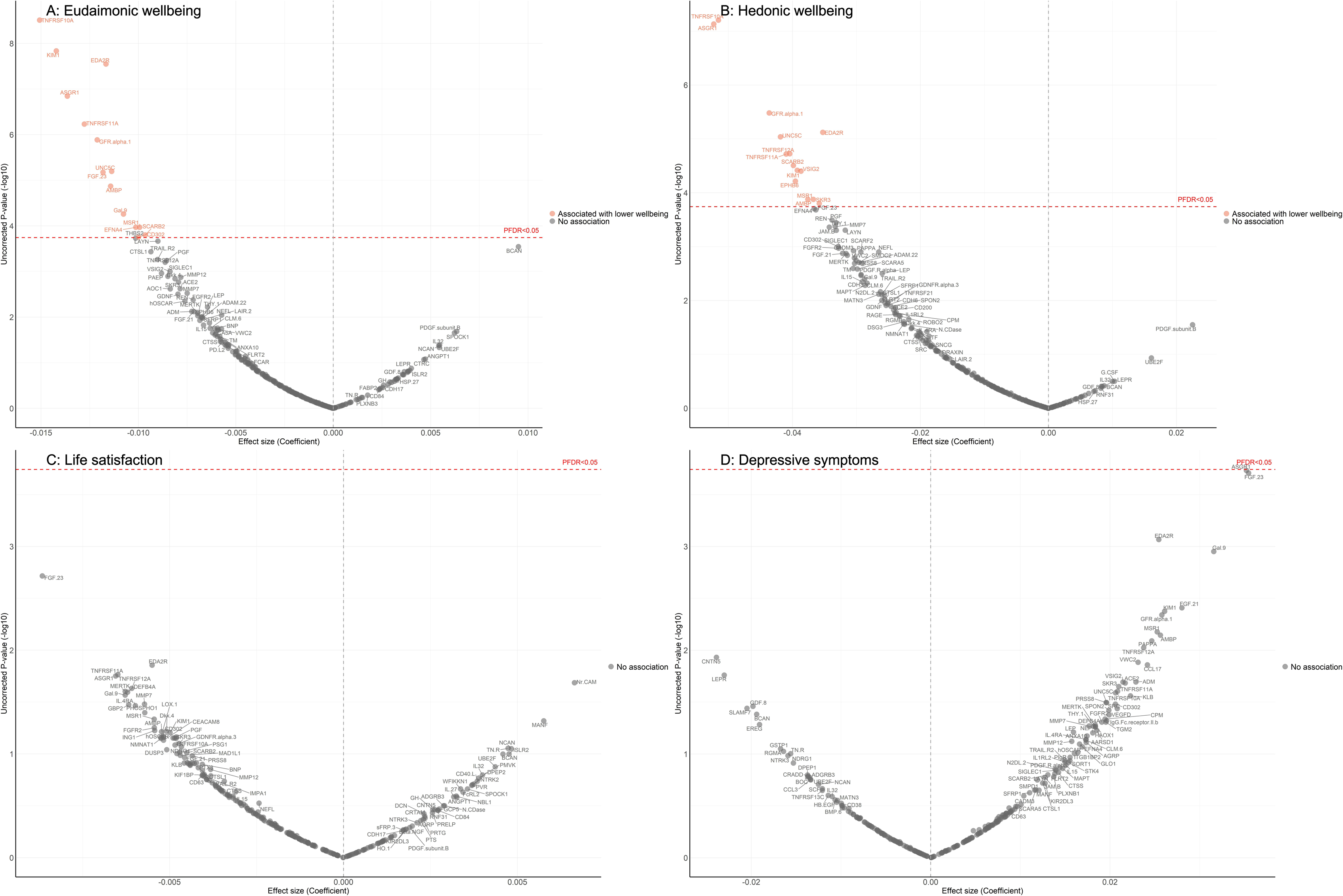
Volcano plots visualizing protein concentration associated with cross-sectional eudaimonic wellbeing, hedonic wellbeing, depressive symptoms, and life satisfaction at protein measurement from linear regressions. Linear regression models for protein-wellbeing associations at wave 4. Models adjusted for age, sex, wealth quintile, ethnicity, smoking status, arthritis, cancer, stroke, chronic lung disease, diabetes, cardiovascular diseases.

Further adjustment for body mass index (BMI) showed that several cross-sectional protein associations with eudaimonic wellbeing persisted (TNFRSF10A, KIM1 (also known as HAVCR1), EDA2R, ASGR1, TNFRSF11A, GFR.alpha.1 (also known as GFRA1), UNC5C, FGF.23) (Supplementary Figure 5A; Supplementary Table 11), and with hedonic wellbeing (TNFRSF10A, ASGR1, GFR.alpha.1, EDA2R, UNC5C, TNFRSF12A, TNFRSF11A, SCARB2, VSIG2, KIM1, EPHB6) (Supplementary Figure 5B; Supplementary Table 12). For life satisfaction (Supplementary Figure 5C; Supplementary Table 13) and depressive symptoms (Supplementary Figure 5D; Supplementary Table 14).

Mutually adjusting for eudaimonic and hedonic wellbeing did not show any significant associations (all P_FDR_ for interaction > 0.05). No sex difference in any of the cross-sectional protein-wellbeing associations was observed (all P_FDR_ for interaction > 0.05) (Supplementary Table 15-18).

### Analysis 2: Longitudinal associations between protein measures at wave 4 and subsequent wellbeing measures over the next 15 years

In the longitudinal analyses from wave 4 (in 2008) to wave 10 (in 2023), the minimally adjusted results from the mixed-effect linear regression models (LMM) linking protein abundance to the trajectory of each wellbeing domain are provided in Supplementary Figure 6, and in Supplementary Table 19-22. After full adjustments, higher concentrations of seven proteins (EDA2R, DCN, CD38, BNP, MERTK, MMP12, CTSL1) at wave 4 were associated with greater decline in levels of eudaimonic wellbeing in the subsequent years after protein measurement (Figure 2A; Supplementary Table 23), while higher concentrations of two proteins at wave 4 were associated with increasing levels of eudaimonic wellbeing (ECE1, DEFB4A). Higher concentrations of eight proteins (DCN, MMP12, EDA2R, TNFRSF13B, SPON2, DRAXIN, MERTK, BNP) were associated with greater decline in levels of hedonic wellbeing (Figure 2B; Supplementary Table 24). Two proteins (KIM1, EDA2R) were associated with greater decline in levels of life satisfaction (Figure 2C; Supplementary Table 25) (all P_FDR_ < 0.05), while one protein (ECE1) was associated with a slower rate of decline in life satisfaction. No protein was associated with the rate of change in the number of depressive symptoms (Figure 2D; Supplementary Table 26), and no protein was associated with the rate of change in the odds of depression, which was modelled using mixed-effect logistic regressions (MLR) (Supplementary Figure 4B; Supplementary Table 27).

**Figure 2.**
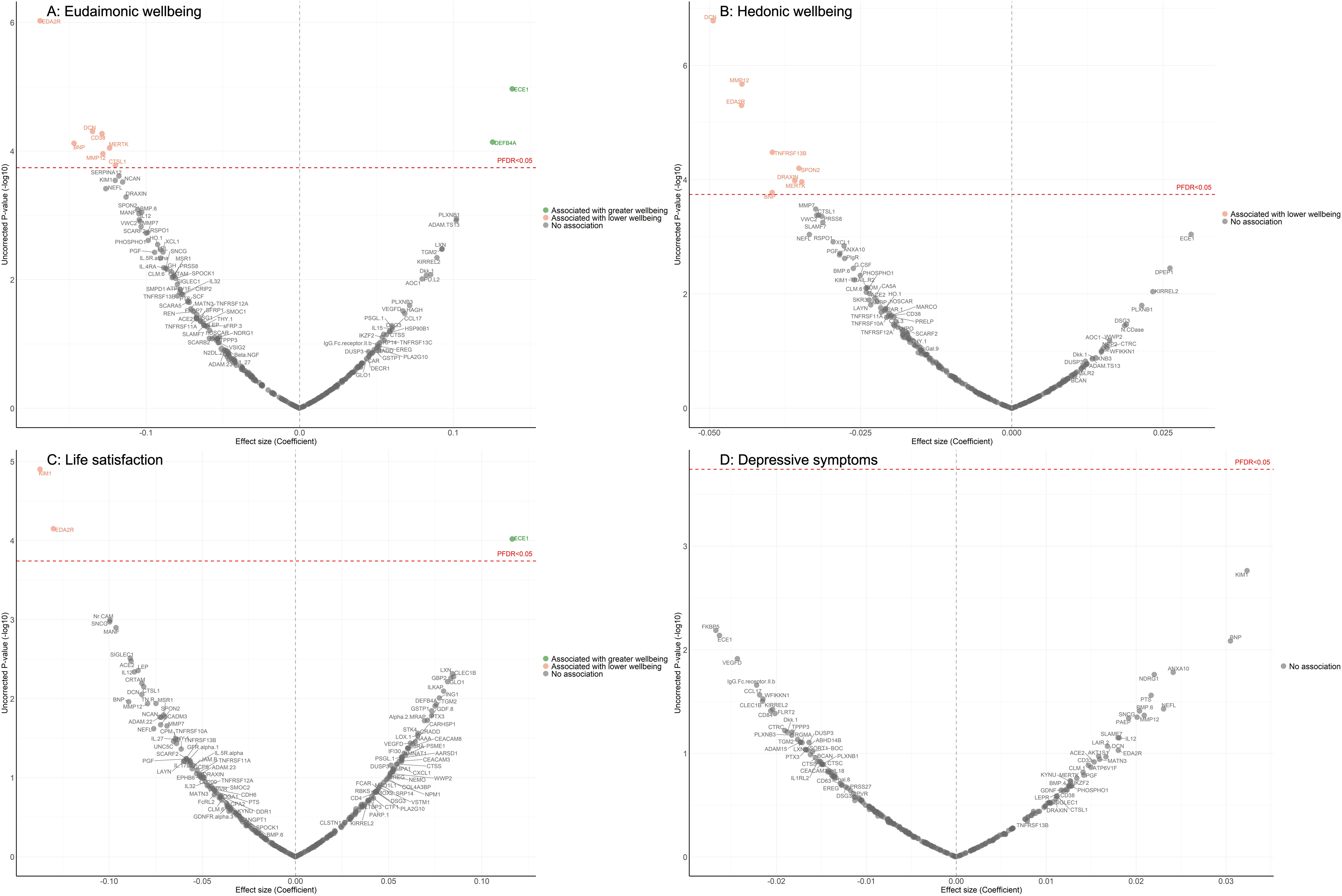
Volcano plots visualizing protein concentration associated with eudaimonic wellbeing, hedonic wellbeing, depressive symptoms, and life satisfaction after protein measurement from linear mixed effect regressions. Linear mixed effect regression models for protein-wellbeing associations from wave 4 to wave 10. Models allowed for protein interacting with time since protein measurements, and adjusted for baseline age, sex, wealth quintile, ethnicity, smoking status, arthritis, cancer, stroke, chronic lung disease, diabetes, cardiovascular diseases.

Further adjustment for BMI largely confirmed the findings described above (Supplementary Table 28-31), with only the association between higher concentrations of BNP and the estimated trajectory of hedonic wellbeing showing attenuation (Supplementary Figure 7B).

For eudaimonic wellbeing, after further adjustment for hedonic wellbeing, the association between higher concentrations of DEFB4A and the rate of change remained significant, showing a positive effect on eudaimonic wellbeing over the 15-year follow-up (coefficient (β) [se] = 0.135 [0.029], P_FDR_ = 0.001). This indicates a distinct and independent relationship eudaimonic wellbeing.

Some notable sex differences were observed (Supplementary Table 32-35). Higher XCL1 concentration was associated with greater decline in levels of eudaimonic wellbeing in males (P_FDR_ for interaction = 0.004; β [se] = -0.24 [0.046], P_FDR_ = 3.32×10^-5^), while it was not significantly associated in females (β [se] = 0.003 [0.041], P_FDR_ = 1.00). This finding of moderation in the longitudinal XCL1-wellbeing association was not attributed to sex differences in levels of XCL1 expression at baseline (p = 0.93).

For hedonic wellbeing, higher concentration of SLAMF7 was also associated with greater decline in levels of hedonic wellbeing in males (P_FDR_ for interaction = 0.02; β [se] = -0.072 [0.014], (P_FDR_ = 6.42×10^-5^), but not in females (β [se] = 0.0007 [0.012], P_FDR_ = 1.00). Testing for differences in protein expression by sex revealed a significant sex difference (p = 0.001), with higher SLAMF7 expression observed in males compared to females.

Internal validation using cluster-level 5-fold cross-validation was applied to evaluate the robustness of protein-wellbeing associations derived from the linear mixed-effects models (Supplementary Tables 36-39). Proteins that were statistically significant in the main models (PFDR < 0.05) were consistently among the strongest contributors to prediction. Cross-validation also indicated, however, that predictive accuracy benefited from the combined contribution of a broader set of proteins, particularly for eudaimonic and hedonic wellbeing, consistent with small, distributed effects across the proteome. Cross-validated RMSE values were highly stable across proteins (range: 6.559-6.593 for eudaimonic wellbeing, 1.735-1.744 for hedonic wellbeing, 6.005-6.014 for life satisfaction, and 1.718-1.722 for depressive symptoms), demonstrating consistent predictive performance across folds. While this supports the robustness of estimates within the sample, external validation will be required to confirm generalizability to independent populations. Overall, these findings suggest that although certain proteins show robust statistical associations, their isolated predictive utility at the individual level remains limited, with more meaningful prediction emerging from distributed multivariate patterns.

### Analysis 3: Longitudinal associations between protein measures at wave 4 and wellbeing in the previous 6 years

Participants scores across the measures of wellbeing were modelled over the six years before protein measurement, spanning wave 1 (in 2002) to wave 4 (in 2008), except for life satisfaction which was modelled from wave 2 (in 2004) to wave 4 (in 2008). Minimally adjusted findings for each wellbeing domain are shown in Supplementary Figure 8 and in Supplementary Table 40-43.

Fully adjusted results from LMM indicate that higher concentrations of four proteins (NEFL, KIM1, EDA2R, GFR.alpha.1) were associated with greater decline in levels of eudaimonic wellbeing in the preceding years (Figure 3A; Supplementary Table 44), while 12 proteins (NEFL, SMOC2, GFR.alpha.1, DCN, SCARF2, TNFRSF12A, EDA2R, SCARA5, PDGF.R.alpha, MATN3, SCARB2, KIM1) were associated with greater decline in hedonic wellbeing (Figure 3B; Supplementary Table 45) (all P_FDR_ < 0.05). Higher concentration of one protein (PSG1) was associated with greater decline in life satisfaction (Figure 3C; Supplementary Table 46). No protein was associated with the level of depressive symptoms (Figure 3D; Supplementary Table 47); or with the binary outcome of depression (Supplementary Figure 4C; Supplementary Table 48). Further adjustment for BMI, based on the fully adjusted models, yielded results largely consistent with those described above, with higher concentrations of KIM1 showing an additional association with lower hedonic wellbeing (Supplementary Figure 9) (Supplementary Table 49-52).

**Figure 3.**
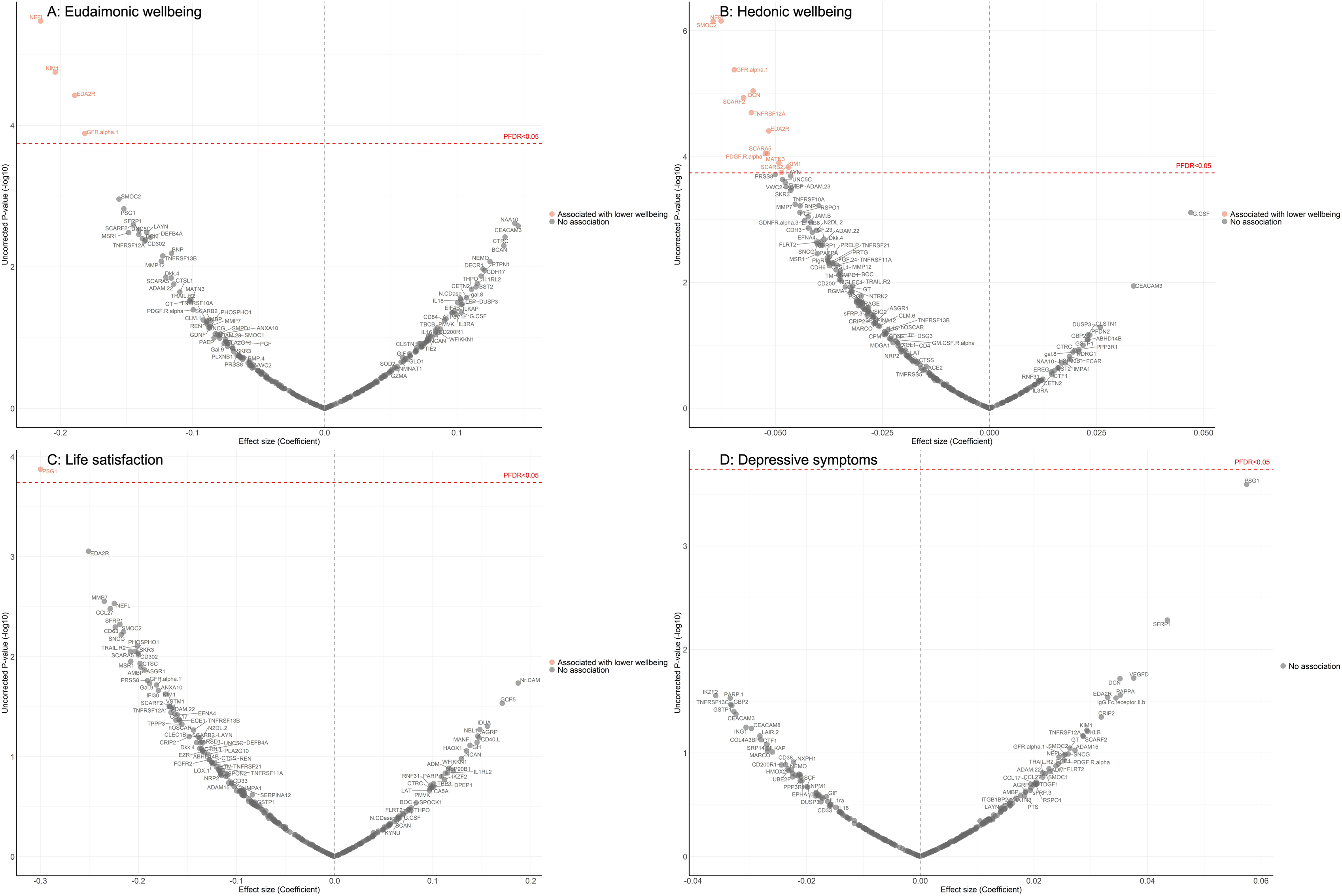
Volcano plots visualizing protein concentration associated with eudaimonic wellbeing, hedonic wellbeing, depressive symptoms, and life satisfaction before protein measurement from linear mixed effect regressions. Linear mixed effect regression models for protein-wellbeing associations from wave 1 to wave 4 (except for life satisfaction, while was from wave 2 to wave 4). Models allowed for protein interacting with time since protein measurements, and adjusted for baseline age, sex, wealth quintile, ethnicity, smoking status, arthritis, cancer, stroke, chronic lung disease, diabetes, cardiovascular diseases.

No significant associations were observed after mutually adjusting for eudaimonic and hedonic wellbeing.

No differences by sex were observed for the longitudinal associations between protein concentration and wellbeing measured in the years before protein assays (all P_FDR_ for interaction > 0.05) (Supplementary Table 53-56).

Cluster-level 5-fold cross-validation produced results consistent with the main analyses (Supplementary Tables 57–60), supporting the robustness of significant protein–wellbeing associations. For hedonic wellbeing, predictive accuracy benefited from the contribution of a broader set of proteins. Cross-validated RMSE values were highly stable across proteins (range: 6.287-6.319 for eudaimonic wellbeing, 1.644-1.652 for hedonic wellbeing, 6.123-6.133 for life satisfaction, 1.809-1.814 for depressive symptoms), indicating consistent predictive performance across validation folds.

Figure 4 summarizes the effect sizes of the cross-sectional and longitudinal associations - both before and after protein measurement - between protein concentrations and the various dimensions of wellbeing, with the significance level of the associations being indicated by P_FDR_.

**Figure 4.**
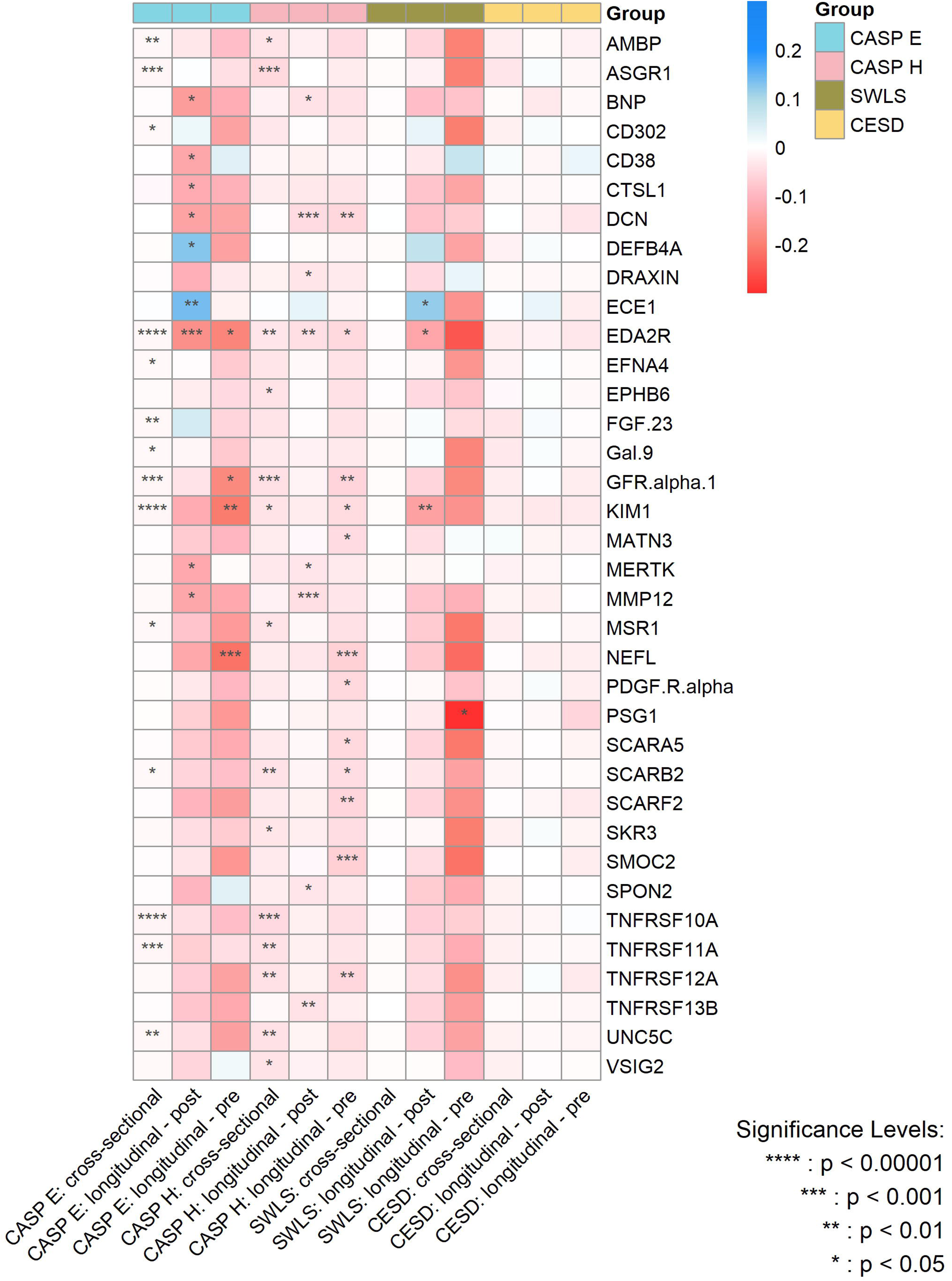
Heatmap for the magnitude of associations between significant proteins and various wellbeing domains and depressive symptoms from cross-sectional and longitudinal analyses after multiple adjustments. CASP E, Control, Autonomy, Self-Realization and Pleasure eudaimonic wellbeing; CASP H, Control, Autonomy, Self-Realization and Pleasure hedonic wellbeing; SWLS, Diener Life Satisfaction scale; CESD, Center for Epidemiological Studies-Depression scale. P values are after false discovery rate (FDR) correction.

### Analysis 4: Enrichment network analysis of the identified proteins

Enrichment network analysis suggested several enriched pathways for all identified proteins in the observed cross-sectional and longitudinal associations with eudaimonic wellbeing (Figure 5; Supplementary Table 61), hedonic wellbeing (Figure 6; Supplementary Table 62), and life satisfaction (Figure 7; Supplementary Table 63).

**Figure 5.**
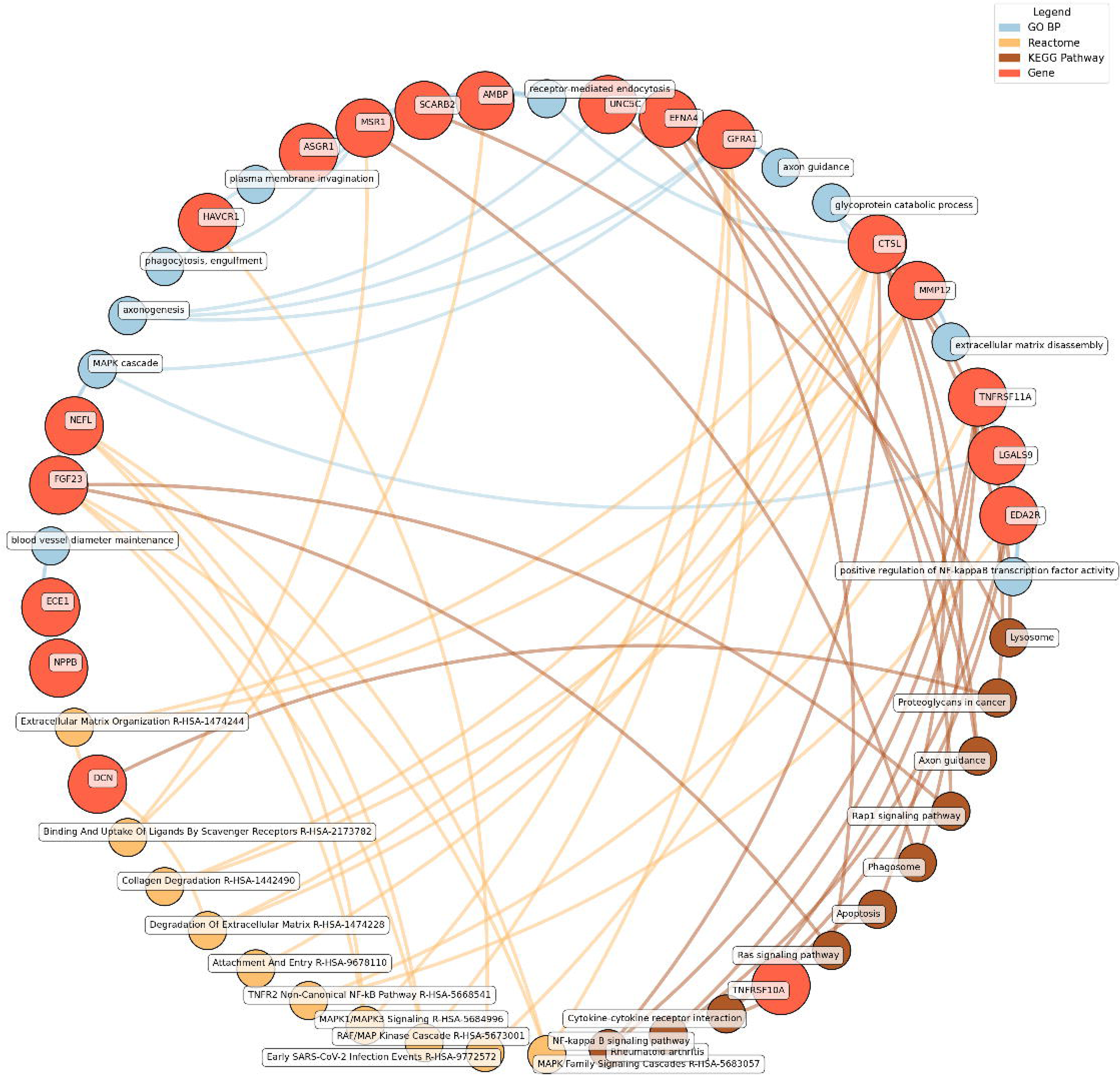
Network analysis for pathways enriched in proteins significantly associated with eudaimonic wellbeing. GO BP, Gene ontology – biological processes; Reactome, a database of reactions, pathways and biological processes; KEGG, Kyoto Encyclopedia of Genes and Genomes. The ‘node’ indicates the gene name corresponding to the protein, or the biological term extracted from each bioinformatics library, the ‘edge’ connects protein to their enriched term.

**Figure 6.**
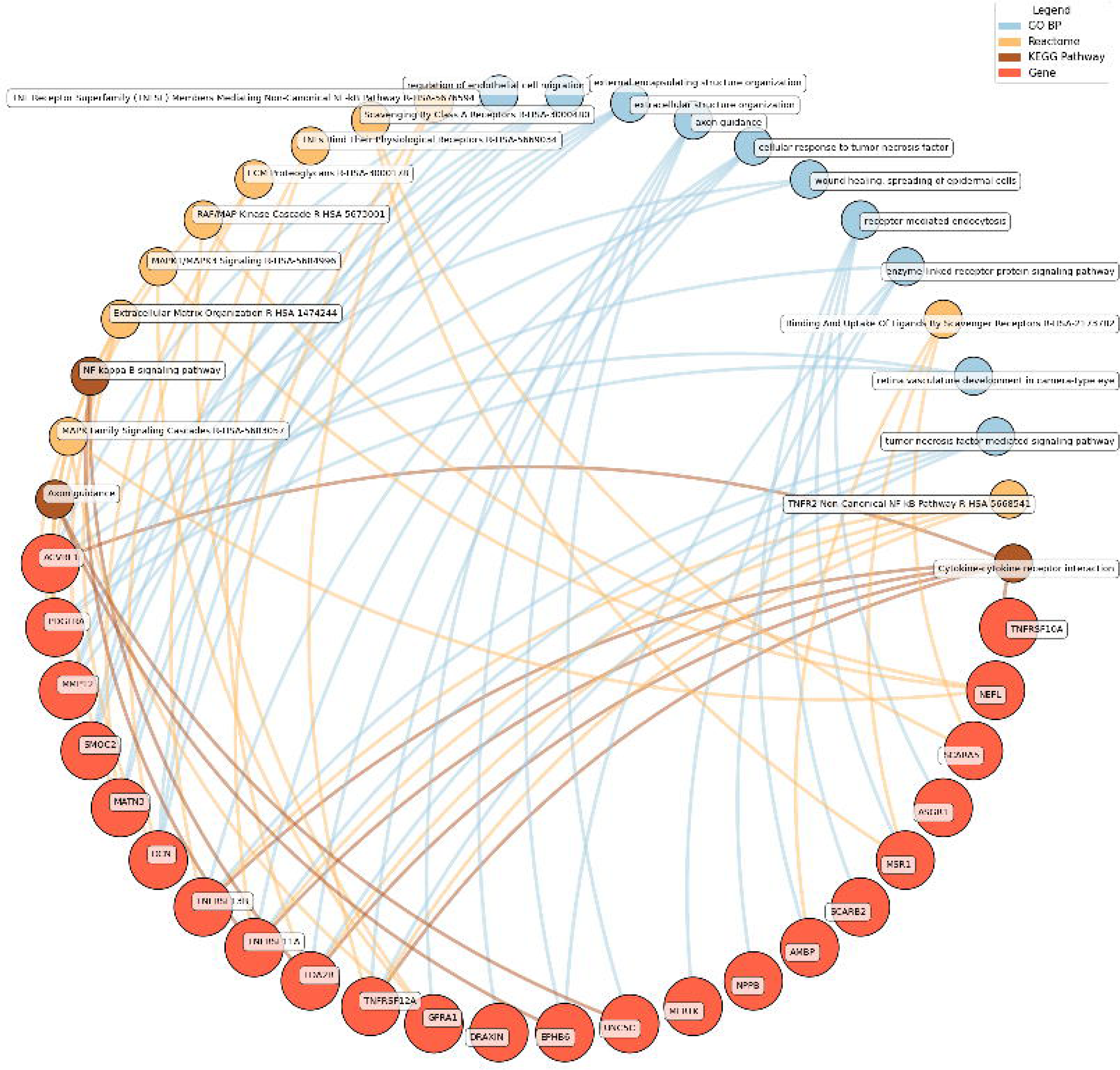
Network analysis for pathways enriched in proteins significantly associated with hedonic wellbeing. GO BP, Gene ontology – biological processes; Reactome, a database of reactions, pathways and biological processes; KEGG, Kyoto Encyclopedia of Genes and Genomes. The ‘node’ indicates the gene name corresponding to the protein, or the biological term extracted from each bioinformatics library, the ‘edge’ connects protein to their enriched term.

**Figure 7.**
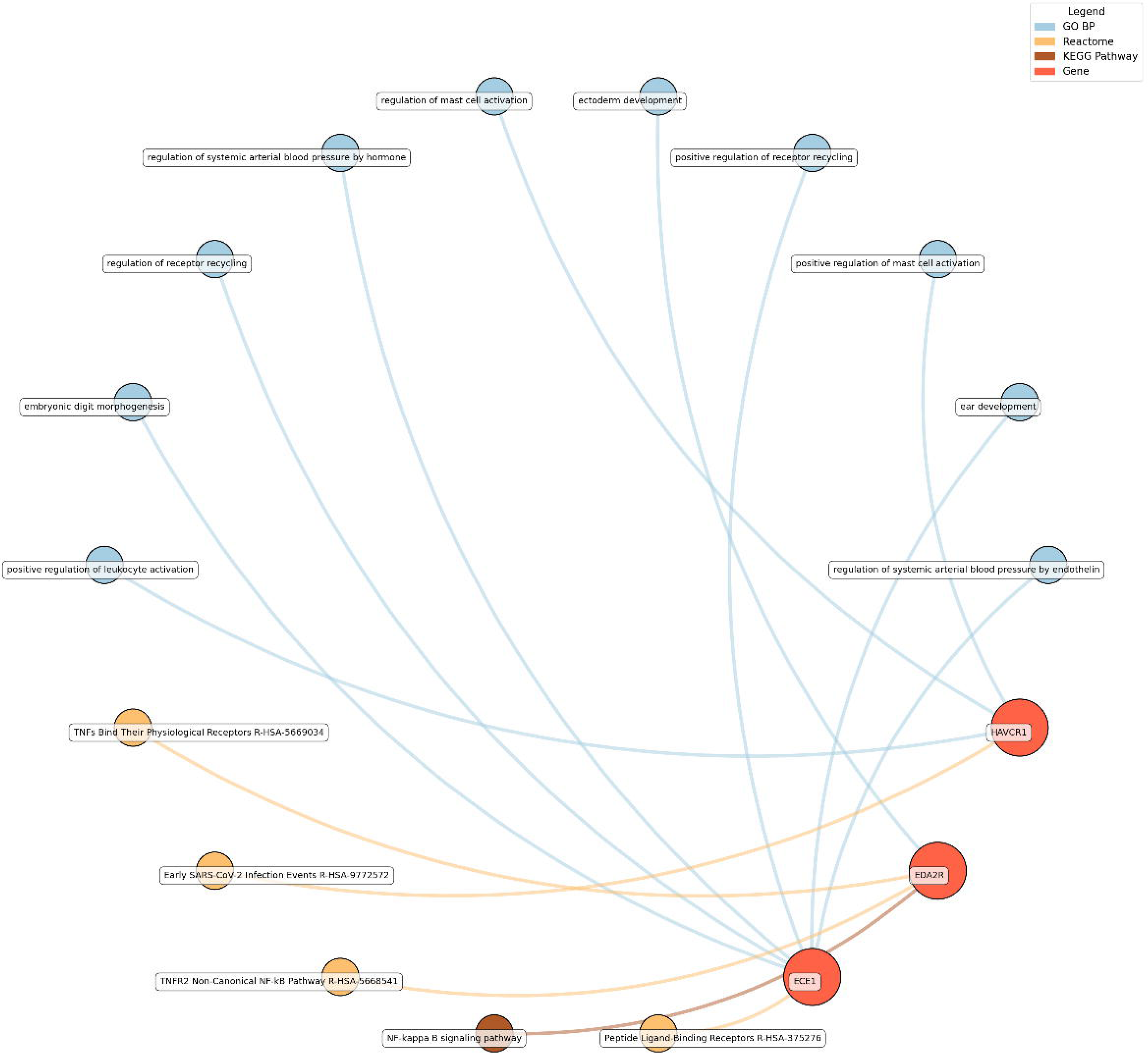
Network analysis for pathways enriched in proteins significantly associated with life satisfaction. GO BP, Gene ontology – biological processes; Reactome, a database of reactions, pathways and biological processes; KEGG, Kyoto Encyclopedia of Genes and Genomes. The ‘node’ indicates the gene name corresponding to the protein, or the biological term extracted from each bioinformatics library, the ‘edge’ connects protein to their enriched term.

Significant pathways enriched for low eudaimonic wellbeing included NF-κB signaling, cytokine-cytokine receptor interactions, and MAPK family signaling cascades, along with extracellular matrix organization and axon guidance. Proteins such as EDA2R, LGALS9, and TNFRSF11A were linked to NF-κB transcriptional activity, while MMP12 and CTSL contributed to extracellular matrix disassembly and glycoprotein catabolism.

Low hedonic wellbeing was particularly associated with TNF-mediated pathways, with proteins such as TNFRSF13B and EDA2R participating in TNF signaling. Extracellular structure organization and axon guidance were also enriched, with contributions from proteins such as DCN, MMP12, and GFRA1.

For life satisfaction, pathways related to systemic arterial blood pressure regulation, receptor recycling, and ectoderm development were prominent, with proteins such as ECE1 and KIM1 (also known as HAVCR1) playing key roles.

Eudaimonic, hedonic, and life satisfaction wellbeing share several biological pathways and processes, reflecting overlapping molecular mechanisms (which is unsurprising given the substantial overlap in protein correlates of each type of wellbeing, e.g., among the higher concentrations of 14 proteins linked to low hedonic wellbeing or low eudaimonic wellbeing, 8 were shared). Common shared pathways include the NF-κB signaling pathway, which is central to inflammatory and immune responses, and the cytokine-cytokine receptor interaction pathway, involved in intercellular communication during immune processes.

Additionally, the MAPK signaling cascades, crucial for cellular responses to environmental stimuli, are shared between eudaimonic and hedonic wellbeing.

## DISCUSSION

This study explored 276 proteomic markers associated with various dimensions of psychological wellbeing, employing a cross-sectional and longitudinal study design with a data-driven approach. Higher concentrations of two plasma proteins at wave 4, DEFB4A and ECE1, were associated with positive eudaimonic wellbeing over the 15-year period from wave 4 to wave 10, while ECE1 was also linked to better life satisfaction. Furthermore, higher concentrations of DEFB4A were associated with higher levels of eudaimonic wellbeing over time, even after accounting for concurrent levels of hedonic wellbeing, suggesting a distinct and independent relationship. In contrast, a broader range of proteins were identified as being associated with lower levels of wellbeing subsequent to protein measurement, with shared patterns observed across both the eudaimonic and hedonic wellbeing domains. The enrichment network analyses suggested that some of these proteins are involved in cytokine-receptor interactions, TNF and NF-κB signaling, as well as MAPK family signaling cascades underscoring the pivotal role of immune and inflammatory responses in relation to overall psychological wellbeing. Proteins involved in axon guidance appear to play an important role in wellbeing, too. Lastly, XCL1, a chemokine, and SLAMF7, involved in the regulation and interconnection of both innate and adaptive immune response, displayed significant sex differences in their associations with estimated declines in levels of wellbeing, with the detrimental effects being primarily observed in men.

A meta-analysis examining biomarkers and wellbeing found moderate negative associations between C-Reactive Protein (CRP), interleukin-6 (IL-6), and fibrinogen with wellbeing.[4] An earlier ELSA study indicated that increases in eudaimonic wellbeing was associated with decreased levels of CRP and fibrinogen.[15] Furthermore, the meta-analysis revealed that TNF-α was either negatively associated or had no significant correlation with wellbeing, suggesting a possible adverse connection between inflammatory markers and general wellbeing.[4] TNF-α is a pro-inflammatory cytokine crucial for immune regulation and inflammatory responses.[16] Elevated levels of TNF-α have been associated with various health problems, including depression,[17] fatigue,[18] and cognitive impairment in Alzheimer’s disease.[19] These findings suggest that chronic inflammation, as indicated by elevated TNF-α and other inflammatory markers, may play a role in impairing mental and physical wellbeing. Indeed, in the current study, several proteins associated with low eudaimonic and hedonic wellbeing were found to be involved in pathways responding to TNF-α and IL-1. These findings pinpoint the biological cascade through which these inflammatory markers may exert their influence on wellbeing via complex immune responses, further emphasizing the intricate relationship between inflammation and mental health.

The proteins we found to be longitudinally associated with positive wellbeing, DEFB4A and ECE1, are implicated in biological mechanisms that support immune function[20] and vascular health,[21] respectively. DEFB4A belongs to the defensin family, a group of cytotoxic peptides secreted by neutrophils that play crucial roles in the innate immune defense against microbial infections.[20, 22] By enhancing the body’s defense against infections and regulating inflammation, DEFB4A may help maintain a balanced immune response, potentially reducing chronic inflammation.[20] Through its antimicrobial and anti-inflammatory properties, DEFB4A may contribute to overall health and positive wellbeing. ECE1, on the other hand, supports vascular health by regulating endothelin-1 levels, which are critical for maintaining blood vessel tone and blood pressure.[21] Adequate vascular function ensures efficient oxygen and nutrient delivery, supporting energy levels and reducing cardiovascular risks associated with stress,[23] thereby promoting positive wellbeing.[24] Collectively, these mechanisms suggest that DEFB4A and ECE1 may contribute to greater wellbeing over time among older adults by enhancing immune resilience, reducing inflammation, and promoting cardiovascular health. Further research is needed to elucidate the biological pathways through which these proteins contribute to positive wellbeing. Additionally, the distinct association between DEFB4A and positive eudaimonic wellbeing, after accounting for hedonic wellbeing, warrants further investigation.

Pathways involving cytokines were enriched for several proteins in relation to low wellbeing. Cytokines have important role in regulating immune and inflammatory responses,[25] which are crucial in maintaining homeostasis as well as wellbeing.[25] Accumulating evidence suggests that pro-inflammatory cytokines play a role in the development of atherosclerosis, major depression, and related conditions such as visceral obesity, metabolic syndrome, and sleep disorders.[25] Cytokines also play a key role in the development of autoimmunity and the progression of autoimmune diseases.[26] During an immune and inflammatory response, the central nervous system and immune system as major adaptive systems of the body, engage in constant communication, a process crucial for maintaining homeostasis.[27] When these systems fail to effectively address and resolve inflammation, particularly chronic inflammation, it can negatively impact an individual’s wellbeing.[25] It is also plausible that subclinical infections contribute to interferon-mediated responses or sickness behaviors that affect mood and alter social motivation.[28] Our analysis using a data-driven approach using largely neuro-relevant protein panels, identified proteins involved in cytokine receptor signaling and response pathways which may be relevant for wellbeing, providing additional markers beyond the focus on TNF-α and interleukins in previous studies. Regarding potential subclinical infections, it is notable that several of the proteins linked to low wellbeing are involved in hepatis virus infection (KIM1, ASGR1) and replication (AMBP, MSR1) as well as other liver-related processes such as hepatic inflammation, cancer, and alcohol-related liver disease (EDA2R, GFRA1, UNC5C, TNFRSF10A, TNFRSF11A).

Moreover, the findings on sex differences in XCL1 in relation to eudaimonic wellbeing and SLAMF7 for hedonic wellbeing over the 15-year period from wave 4 to wave 10 were particularly intriguing, revealing that the potential harmful effects of these proteins on wellbeing among older adults appear to be restricted to men. XCL1 is a pro-inflammatory chemokine that plays a critical role in immune responses by attracting T cells and other immune cells to sites of inflammation.[29] It is primarily produced by activated T cells, dendritic cells, and other immune cells, with its main function being the regulation of lymphocyte trafficking. XCL1 expression is elevated in several infections and autoimmune diseases.[29] SLAMF7 has significant role in modulating the activation and differentiation of a wide variety of immune cells and thus are involved in the regulation and interconnection of both innate and adaptive immune response.[30–32] While sex differences in immune responses have been well-documented, the underlying mechanisms remain unclear.[33] One hypothesis suggests that sex chromosomes may play a role, given that several immune-related genes are encoded on the X chromosome. However, this is unlikely to explain the sex differences observed in our study, as the XCL1 and SLAMF7 encoding gene are both located on Chromosome 1. Sex plays a crucial role in shaping the transcriptomes of immune cells, with aging affecting the immune system differently in males and females.[33] Moreover, sex hormones, such as estradiol (a major form of estrogen), which is a major female steroid hormone, may also contribute to these differences.[33, 34] For example, estradiol has been shown to reduce the production of excessive innate inflammatory cytokines by monocytes and macrophages, offering a potential explanation for sex-based differences in immune responses.[34] By contrast, testosterone can markedly increase the production of SLAMF7 and a related population of pro-inflammatory macrophages.[35] If sex differences in immune responses exist, they could help explain the negative correlation between XCL1 and SLAMF7 with wellbeing in men. In older males, changes in the epigenetic landscape of immune cells may lead to hyperinflammation, while in females, estradiol’s suppression of inflammatory cytokines may offer protection.[33] These distinct mechanisms could be key factors driving sex differences in age-related outcomes, including multi-dimensional wellbeing.

Several proteins involved in neurotrophic signaling such as axon guidance (GFRA1, EFNA4, DRAXIN, EPHB6, and UNC5C) and axonogenesis (GFRA1, EFNA4, and UNC5C) were shown to be significantly associated with lower eudaimonic and hedonic wellbeing. Axon guidance is a critical process in neurodevelopment that directs the growth of neuronal axons to their appropriate targets, ensuring proper neural network formation.[36] The role of axon guidance extends beyond the development phase and influences lifelong neural plasticity and adaptability.[37] Dysregulation of axon guidance mechanisms can lead to neurodevelopmental disorders and late-onset brain conditions, such as neurodegenerative diseases,[37] which significantly impact an individual’s wellbeing. For example, aberrant axon guidance may impair emotional wellbeing by disrupting neural circuits responsible for emotional regulation, increasing susceptibility to psychiatric disorders such as depression.[38] Similarly, it is implicated in neurodegenerative diseases such as Parkinson’s disease, potentially through dopaminergic circuits,[37] which are closely linked with manifestations such as anhedonia and emotional apathy, especially when dopamine levels are low.[4] Understanding these mechanisms may open pathways for developing therapies that support neural regeneration or compensate for lost connectivity, ultimately enhancing wellbeing.

Taken together, our findings delineate a complex biological landscape underlying human psychological wellbeing, revealing some convergent pathways for its eudaimonic and hedonic dimensions. We demonstrate that both forms of wellbeing share a common immunoregulatory foundation. This shared substrate is characterized by enhanced efficiency in innate immune surveillance mechanisms, notably scavenger receptor-mediated endocytosis, which facilitates the clearance of cellular debris and modified lipoproteins.[39, 40] Furthermore, both states are associated with a nuanced regulation of the NF-κB signaling pathway, suggesting a poised, yet well-controlled, inflammatory response capacity that may confer resilience against cellular stress.[10] Beyond immunity, our data implicate shared investments in physiological maintenance, evidenced by enrichment in pathways governing neural circuit integrity (e.g., axon guidance) and the preservation of structural tissue health through extracellular matrix (ECM) organization.

Beyond the shared biology, our analysis uncovered pronounced, functionally distinct biological profiles between eudaimonic and hedonic wellbeing. Eudaimonic wellbeing was uniquely associated with a catabolic phenotype, and this is defined by the upregulation of processes dedicated to the systematic breakdown and turnover of macromolecules and the ECM.[41] In line with prior work linking eudaimonia to enhanced antiviral defences,[10] we found a specific enrichment for the SARS-CoV-2 infection pathway. In contrast, hedonic wellbeing was characterized by a pronounced association with the TNF superfamily signalling network, indicates a connection to the body’s primary inflammatory signalling axis.[42] This aligns with previous gene expression studies linking positive affect to an upregulation of pro-inflammatory transcripts.[10] Concurrently, hedonic wellbeing was also distinguished by a signature of anabolic reconstruction, including pathways involved in vasculogenesis, wound healing, and synthesis and organization of ECM proteoglycans. This juxtaposition of inflammatory signalling and tissue-building processes suggests a physiological state geared towards reactive repair and response to environmental stimuli or perceived needs. This biological divergence provides a novel framework for understanding longstanding psychological theories. Eudaimonic wellbeing, which emphasizes systemic maintenance, turnover, and antiviral defence, resonates with models that conceptualize eudaimonia as a state of deep engagement and purposeful striving, necessitating efficient physiological upkeep and long-term defence investment.[6] Conversely, hedonic wellbeing, characterized by acute TNF-driven signalling and reactive tissue repair, mirrors a more reactive pursuit of rewards, is linked to the body’s reactive communication systems and processes that build and repair tissues in response to external stimuli or needs.

While numerous studies have explored the associations between biomarkers and wellbeing, they often face limitations such as examining only a small subset of biomarkers and being hypothesis-driven rather than data-driven in their approach, with the latter being comparatively rare. A recent data-driven study using proteomics data from the UK Biobank revealed significant associations between several proteins and the risk of various age-related diseases.[43] Specifically, the study found that several proteins were associated with the risk of mood disorders.[43] However, there was no examination of wellbeing outcomes. The Malmö Diet and Cancer Cardiovascular Cohort study found 34 plasma proteins to be associated with poor self-rated health using a panel of cardiovascular risk proteins in 4,521 participants without cardiovascular diseases.[44] This study found that two proteins, namely leptin and CCL20, showed significant relationships even after adjustments for multiple risk factors, and that these proteins could potentially play a role in the relationship between self-rated health and cardiovascular diseases beyond traditional risk factors.[44] It was somewhat surprising that our study did not find any protein associated with the number of depressive symptoms and the binary outcome of depression longitudinally, despite accumulating biomarker literature showing such relationships.[45] Several factors may help explain this null finding. First, depressive symptoms in ELSA were assessed using the 8-item CESD scale, which serves as a brief symptom screener rather than a diagnostic instrument. The distribution of scores in this general population sample of older adults may not have captured sufficient variability or severity to detect proteomic associations that typically emerge in clinical or high-symptom samples. Second, the proteomics platforms assayed in this study may not have included key biomarkers specifically relevant to depression, limiting coverage of relevant biological pathways. Third, wellbeing and depression, while related, are not merely opposite ends of the same continuum; they may have partially distinct biological underpinnings, such that proteomic associations with positive psychological states do not necessarily mirror those with depressive symptoms.

To the best of our knowledge, this is the first study to assess the relationships between proteomics markers and various dimensions of positive psychological wellbeing in older adults, by undertaking a proteome-wide data-driven approach. This provided novel and nuanced evidence for biological pathways involved in psychological wellbeing. Noteworthy strengths underpin this study, including its expansive sample size in a nationally representative cohort of older adults. Additionally, this study conducted cross-sectional and longitudinal assessments of the relationships between protein markers and wellbeing measures both before and after protein measurement, affording a 20-year span of wellbeing measures, thereby enhancing the robustness of the findings. The inclusion of multi-dimensional wellbeing was another major strength.

However, some limitations should be acknowledged. Our proteomic assays covered only a small fraction of the roughly 20,000 proteins catalogued by the Human Proteome Project.[46] That said, the selected protein panels were primarily focused on neurological and cardiovascular processes, which may be especially relevant when examining wellbeing trajectories. A study using UK Biobank data explored the role of brain health in mediating the relationship between physical and mental health, focusing on depression and anxiety. The results showed that brain health had a significant mediating effect, particularly on the musculoskeletal and immune systems, which themselves exerted strong direct impacts on mental health outcomes.[47] Additionally, life satisfaction in the ELSA study was measured over a shorter period (starting from wave 2 in 2004), so some of the associations prior to protein measurement may be less robust, as they estimate changes over only approximately four years. Furthermore, the absence of longitudinal proteomic profiling measurements in our study, as is the case in most studies in proteomics, presents another notable constraint. Because data were collected at a single time point, we cannot determine the temporal ordering of associations. Thus, it remains equally plausible that variation in proteomic profiles influences wellbeing or that wellbeing shapes proteomic patterns through behavioral and physiological processes. Future studies employing repeated or experimental designs will be essential to disentangle these causal pathways. Additionally, despite examining the long temporal associations over 20 years, firmer causal evidence using causal inference methods is still needed, such as using Mendelian randomization (MR). We were unable to perform MR in our study given the lack of appropriate Genome-Wide Association Studies related to specific wellbeing domains. Lastly, we lacked a suitable external cohort with comparable measures of circulating proteins and repeated assessments of wellbeing to validate our findings. This limits our ability to directly test the reproducibility of the associations we observed in an independent sample. Future studies that include both proteomic and longitudinal wellbeing data in independent populations will therefore be essential to confirm the robustness and external validity of our results.

In conclusion, this study expands on previous literature on the biological markers involved in positive and negative wellbeing trajectories, and uncovered novel biological pathways related to different wellbeing domains. Consequently, these markers may serve as potential indicators that help bridge biological, psychological, and social factors in comprehending wellbeing, guiding interventions aimed at promoting health and wellbeing. Notably, cytokines’ involvement in immune systems appears to be particularly relevant in relation to wellbeing trajectories in older adults. Future studies are needed to better understand the involvement of cytokine dysfunction and to explore the biological underpinnings of the inflammatory and immune systems, and neurotrophic signaling in psychological wellbeing.

## ONLINE METHODS

### Study population

The study utilized data from ELSA, a nationally representative longitudinal study of approximately 12,000 individuals aged 50 and above, along with their partners, residing in private households in England.[14] Commencing with its inaugural wave of data collection in 2002/2003, ELSA employed a combination of in-person interviews and self-completion paper questionnaires with a 2-year follow-up interval. The blood samples collected during the wave 4 nurse visit in 2008 were employed for proteomic analysis in ELSA. Among those with a blood sample, participants who died within two years following the wave 4 nurse visit or who were lost to follow-up (defined as non-participation in at least two consecutive waves) were excluded based on predetermined criteria. The current study encompassed a final combined dataset of 3,262 participants with proteomics assayed (Supplementary Figure 1).

ELSA was approved by the London Multi-center Research Ethics Committee (MREC/01/2/91). Informed consent was obtained from all participants or their guardians.

### Wellbeing indicators

***Hedonic wellbeing*** was assessed using the pleasure subscale of the CASP quality of life questionnaire, which was developed for older people.[48] This four-item pleasure subscale, gauges enjoyment of life through statements such as “I enjoy the things I do”, with response options ranging from “Never” to “Often”. Responses were summed to yield total enjoyment of life score (ranged 0 to 12), with higher scores reflecting greater enjoyment.

***Eudaimonic wellbeing*** was assessed using the remaining 15 items of the CASP quality of life questionnaire.[48] Scores were summed (ranged 0 to 42), with higher scores reflecting greater enjoyment. It has been found that this measure of eudaimonic wellbeing is associated with survival after demographic, health status, health-related behaviors and depression have been taken into account.[49]

***Life satisfaction (or evaluative wellbeing)*** was measured using the Diener Life Satisfaction scale (SWLS).[50] This scale comprises five items assessing overall life satisfaction, with response options on a seven-point scale from “Strongly agree” to “Strongly disagree”. Example statements include “In most ways my life is close to my ideal”. Scores were summed (range 0 to 30), with higher scores indicating greater life satisfaction.

Self-reported ***depressive symptoms*** were evaluated using an abbreviated eight-item version of the Center for Epidemiologic Studies Depression (CESD) scale.[51] Participants were asked to indicate, via a binary yes or no response, whether they had experienced symptoms such as restless sleep and unhappiness during the past week. Responses were summed (ranged 0 to 8), with higher scores indicating more depressive symptoms.

We further dichotomized depressive symptoms measured by CESD into the binary outcome of depressed versus not depressed, with a cut-off point at 3, to indicate the possible presence of clinical depression.[52] This definition has been verified through standardized psychiatric interviews conducted with older populations.[53]

All wellbeing indicators were measured at all study waves in ELSA, except for the life satisfaction domain which was measured from wave 2 (in 2004) onwards. For the current study, we used all available measures from wave 1 (in 2002) to wave 10 (in 2023).

### Proteomics data

The Olink™ technology is described in the Supplementary Methods. For the proteomics assays, three Olink™ Target 96 panels were chosen: Cardiovascular II (CVDII), Neurology I (NEUI), and Neurology Exploratory (NEX), comprising 276 distinct proteins. The frozen samples were sent to Olink for aliquoting, plating, and conducting the assays. These assays integrate an inherent quality control, incorporating four internal controls added to all samples, in addition to external controls. Protein concentrations were quantified utilizing Olink’s standardized Normalized Protein eXpression (NPX) values, presented on a Log_2_ scale. The quality control methods employed to examine the proteomics data have been described previously.[54]

### Covariates

Sociodemographic factors, included age (in years), which was calculated using participant’s date of birth and the date of the wave 4 nurse visit, self-reported sex (female vs. male), ethnicity (white versus other ethnic groups), and total non-pension household wealth, which represented the combined value of financial assets, physical possessions, and housing owned by a household (consisting of either a single respondent or a responding couple with any dependents), minus any debts.[55] Continuous wealth was categorized into quintiles. Smoking status was self-reported, categorized into never, current, and former smoker. BMI was calculated using weight [kilograms]/height [meters]^2^, measured during the nurse visit. Participants self-reported information concerning six physician-diagnosed chronic conditions: cancer, diabetes, coronary heart disease, stroke, chronic lung disease, and arthritis.

### Statistical analysis

#### Proteomics data quality control and missing data handling

Before analysis, protein levels were scaled to have a mean of 0 and a standard deviation of 1.[56] All proteins had <6% missingness. A multiple random forest regression imputation was used to impute missing data from the complete dataset,[57] including protein measurements, and covariates. Missing values for the wellbeing outcome variables were not imputed, given that linear mixed-effects models (LMM) can manage missing outcome data through their robust handling of incomplete longitudinal records.[58]

To examine the correlations between wellbeing measures and depressive symptoms by waves, Spearman’s rank correlation test was used.

We assessed the protein-wellbeing associations in three steps: 1) cross-sectional associations between protein and wellbeing at wave 4; 2) longitudinal associations between wellbeing (wave 4 to wave 10) at and after protein measurements at wave 4; and 3) longitudinal associations between wellbeing (wave 1 to wave 4) at and before protein measurements at wave 4. This approach enables us to capture potential time-dependent effects of proteomics measurements on wellbeing over time.

### Analysis 1: Cross-sectional protein-wellbeing associations at wave 4 using linear or logistic regression models

For the cross-sectional linear regression analysis, the estimates from all 30 imputed datasets were pooled. Initially, protein associations with each of the wellbeing domains were assessed in the imputed datasets, with false discovery rate (FDR)-corrected p-values (denoted as P_FDR_) reported. These results were visualized using a volcano plot, with coefficients (effect sizes) displayed on the x-axis, and a significance threshold of 0.05 for P_FDR_ was applied.

Three sets of covariates adjustments were used: Model 1 specification including age as an interaction term, adjusted for sex, ethnicity, and wealth quintile; Model 2 incorporated the adjustments from Model 1, further adjusted for smoking status, and six chronic diseases (cardiovascular diseases, arthritis, cancer, diabetes, chronic lung disease, and stroke); and Model 3 further adjusted for BMI. Estimates from Model 2 were used to report the main findings.

Additionally, for models examining eudaimonic and hedonic wellbeing as outcomes, all covariates were adjusted, with mutual adjustments made for each other.

For assessing the cross-sectional protein-depression association at wave 4, a logistic regression model was utilized, implemented via the ‘glm’ function in R. The same multiple adjustments as described above were applied.

Sex was included as an interaction term with the protein in the linear regression models to explore potential sex differences. If a sex difference was detected in any of the associations, a test for sex differences in protein expression was conducted using the Wilcoxon Rank-Sum Test, given the non-normal distribution of protein expression levels in both sexes, rendering parametric tests unsuitable.[59]

### Analysis 2: Longitudinal associations between protein measures at wave 4 and subsequent wellbeing measures over the next 15 years using mixed-effect linear or logistic regression models

To examine the long-term effects of protein levels on various wellbeing domains, we used linear mixed-effect (LMM) models constructed with the ‘lmer’ function from the ‘lme4’ package in R. These models assessed how proteins measured at wave 4 influenced wellbeing trajectories over the following 15 years. The models included random intercepts and slopes to account for both within-subject variability and the longitudinal structure of the data. Model convergence was checked for all protein-outcome combinations, and residual diagnostics (normality and homoscedasticity) were evaluated to ensure assumptions were met.

Three sets of covariates adjustments were included in the models, akin to the cross-sectional analysis described above, allowing in the fixed part of the model for interactions between protein levels and the time since protein measurement (thereby allowing the rate of change in wellbeing to vary by protein levels). Model 1 adjusted for baseline age, sex, ethnicity, and wealth quintile; Model 2 further adjusted for smoking status, and presence of diagnosed arthritis, cancer, stroke, chronic lung disease, diabetes, and cardiovascular diseases; Model 3 further adjusted for BMI. Additionally, mutual adjustments were made for eudaimonic and hedonic wellbeing in a separate model.

To assess the longitudinal association between protein levels and the binary outcome of depression, we employed mixed-effects logistic regression (MLR) using the ‘glmer’ function in R. The same covariate adjustments as described above were applied as in the LMM. Additional analyses were conducted by including sex as an interaction term with protein (to formally test for sex differences in the protein and wellbeing associations) and time since protein measurement in both the LMM and MLR models. We considered interaction effects to be statistically significant if the p-value after FDR correction was below 0.05.

We further assessed the robustness and predictive performance of the proteome associations using cluster-level 5-fold cross-validation. To preserve the longitudinal structure of the data, folds were defined at the participant level, such that all repeated measurements from a given individual were contained within the same fold.[60] This ensured that the model was always trained and evaluated on independent sets of participants, thereby preventing information leakage across folds For each cross-validation iteration, a LMM was fitted on four folds, including random participant intercepts and adjustment for covariates based on Model 2. The fitted model was then used to generate predictions for the held-out fold. Out-of-sample predictive performance was quantified separately for each protein by pooling all predictions across the five folds and calculating the root mean squared error (RMSE) against observed outcomes.

### Analysis 3: Longitudinal associations between protein measures at wave 4 and wellbeing in the previous 6 years using mixed-effect linear or logistic regression models

We repeated the analytical approach from Analysis 2 to investigate the association between protein levels measured at wave 4 and wellbeing in the preceding 6 years. All models were checked for convergence to ensure appropriate estimation. Since life satisfaction was first assessed in the ELSA study at wave 2 (2004), the associations between protein levels and life satisfaction in the preceding 4 years were modelled.

The same LMM and MLR models were applied, with the same imputation methods for missing proteomics and covariate data, and covariate adjustments and interaction terms with time since protein measurement, similar to analysis 2. Analysis to test for sex difference was similarly conducted.

Similarly, the cluster-level 5-fold cross validation was implemented.

### Analysis 4: Enrichment network analysis of the identified proteins

Enrichment network analysis was performed using EnrichR-KG (https://maayanlab.cloud/enrichr-kg),[61] a knowledge graph that integrates annotated gene sets from multiple biological libraries into a unified network.[62]

An input gene list, comprising significant proteins identified in the cross-sectional and longitudinal analyses (Analyses 1-3, based on Model 2) for each wellbeing domain, was submitted to the Enrichr-KG web server. Enrichment analysis was performed against three core biological databases: 1) The Gene Ontology (GO) database categorizes genes into biological processes, molecular functions, and cellular components, facilitating the understanding of protein function and localization within biological systems;[63] 2) The Kyoto Encyclopedia of Genes and Genomes (KEGG) database offers a collection of pathways and functional annotations, allowing for the identification of biological pathways enriched;[64] and 3) The REACTOME database: a database of reactions, pathways and biological processes, contains curated pathways and reactions involved in various biological processes, enabling the exploration of molecular events and signaling pathways.[65]

Enrichr-KG performs a statistical over-representation analysis (Fisher’s exact test) for the input genes against each gene set in these libraries. To construct the network, the system then queries its underlying graph database, which pre-computes associations from these libraries as “gene-term” relationships, to retrieve the top significantly enriched terms. For our analysis, we extracted the top 10 significantly enriched terms from each of the three libraries based on a p-value threshold of 0.05.

This process based on the gene set libraries from EnrichR-KG[61, 66, 67] generated a bipartite network of nodes and edges, where nodes represent either (1) gene nodes representing the significant proteins from our input list, and (2) term nodes representing the significantly enriched biological pathways from GO, KEGG, and Reactome. Edges connect a gene node to a term node if that gene is a documented member of that gene set.

This integrated network approach allows for the identification of central biological themes (highly connected terms) and key hub genes that are associated with multiple enriched functions, which might be missed when examining libraries in isolation. Thus, this method enhances the discovery of connections between genes and functional terms, revealing clustered biological signatures across multiple datasets that are difficult to detect with traditional list-based enrichment methods.

The node and edge data for this enriched subgraph were exported from Enrichr-KG. Final visualization was performed programmatically using the ‘NetworkX’ library in Python (version 3.11.7).

## Data Availability

The ELSA data is available on the UK Data Service.

## Acknowledgement

We would like to thank the participants in ELSA for their contribution to the research.

The English Longitudinal Study of Ageing is funded by the National Institute on Aging (grant number R01AG17644) and the National Institute for Health and Care Research (198/1074-02). The National Institute of Aging (NIA) (grant Number [R01AG17644]) funded the proteomics data curation in ELSA. J.G. is supported by the NIA (grant Number [R01AG17644]).

The funders had no role in study design; in the collection, analysis and interpretation of data; in the writing of the report; and in the decision to submit the article for publication.

## Author Contribution

JG and AS conceptualized the study. JG led the methodology, with support from SS. JG carried out the formal analysis and data visualization. JG and AS were responsible for the data curation. JG drafted the original manuscript. All authors participated in review and editing. AS supervised the study. AS and PZ were responsible for funding acquisition. All authors accept responsibility for submitting the work for publication.

## Code availability

The codes used for all analyses are available on GitHub repository: https://github.com/jgong94/ELSA_proteomics_wellbeing.

