## Supplementary Figures for "Associations between plasma proteins and psychological wellbeing: evidence from over 20 years of the English Longitudinal Study of Ageing"

**Supplementary Figure 1. Schematic diagram of participant selection in ELSA proteomics study.**

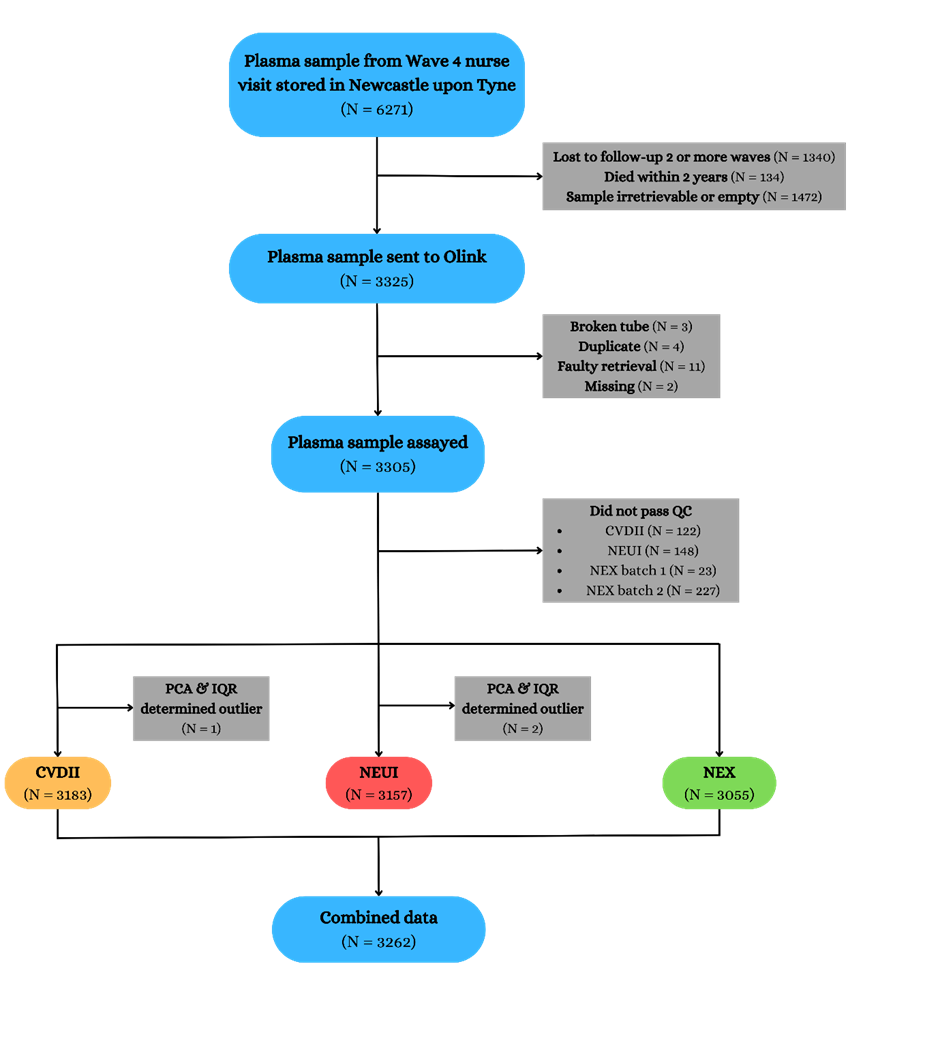

**Supplementary Figure 2. Correlation heatmap for eudaimonic wellbeing, hedonic wellbeing, life satisfaction and depressive symptoms by study wave.**

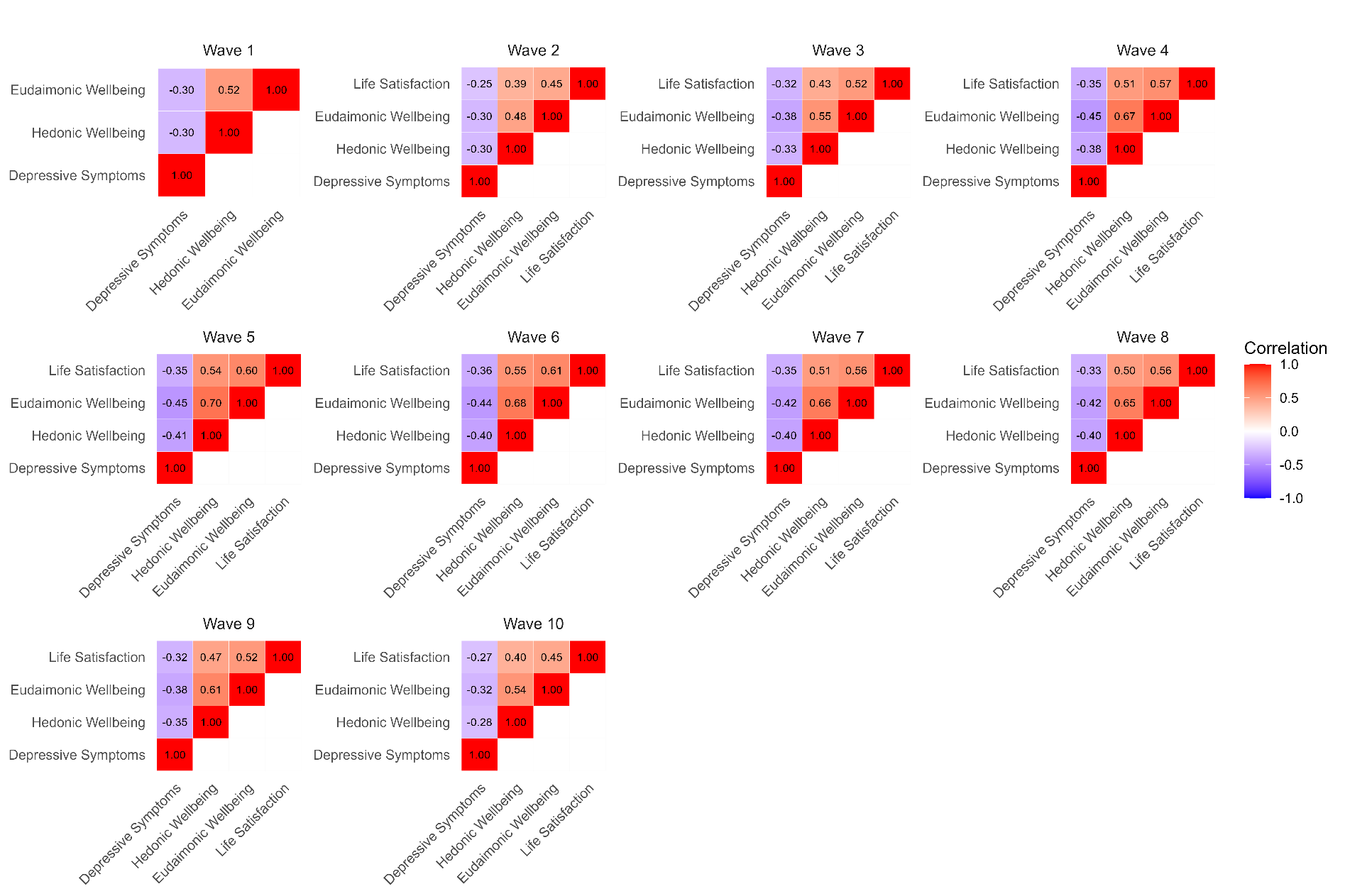

Life satisfaction was not measured in wave 1.

**Supplementary Figure 3. Volcano plots visualizing protein concentration associated with cross-sectional eudaimonic wellbeing, hedonic wellbeing, depressive symptoms, and life satisfaction at protein measurement from linear regressions, with minimal adjustments.**
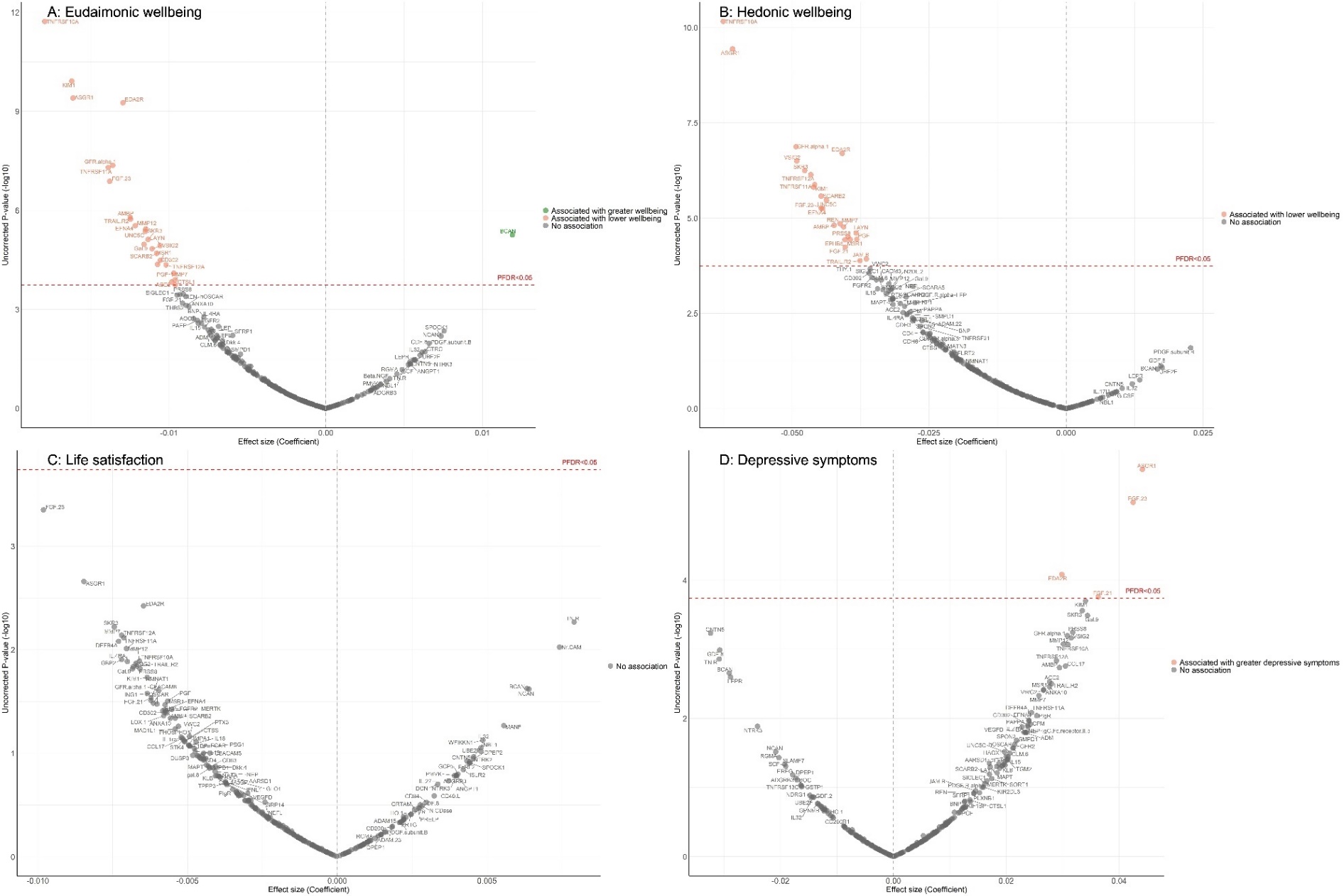

Linear regression models for protein-wellbeing associations at wave 4. Models adjusted for age, sex, wealth quintile, ethnicity.

**Supplementary Figure 4. Volcano plots visualizing protein concentration associated with cross-sectional and longitudinal depression from fully adjusted logistic and mixed-effects logistic regressions.**

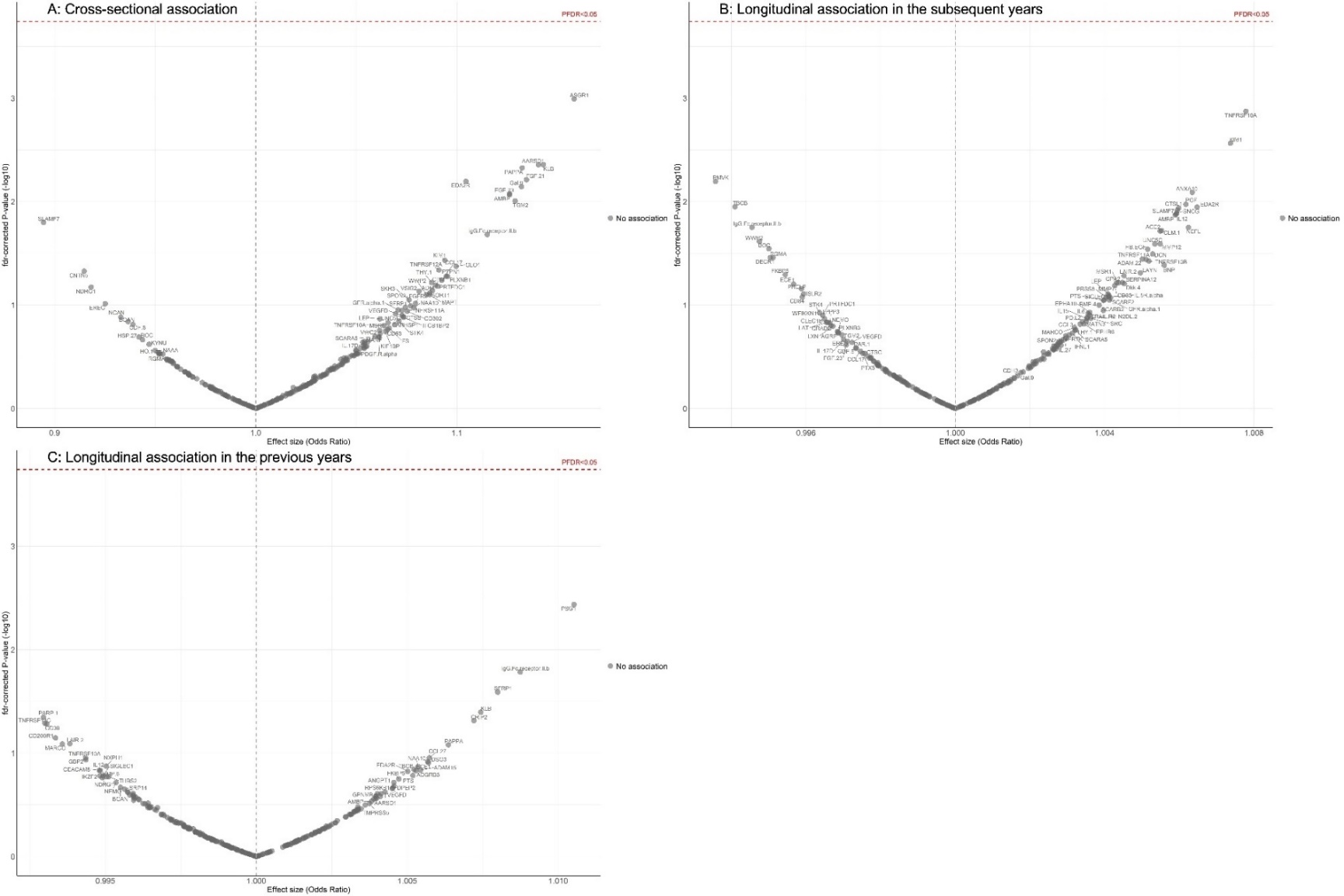

For cross-sectional associations, logistic regression models adjusted for age, sex, wealth quintile, ethnicity, smoking status, arthritis, cancer, stroke, chronic lung disease, diabetes, cardiovascular diseases.

For longitudinal associations, mixed effect logistic regression models allowed for protein interacting with time since protein measurements and age, and adjusted for age, sex, wealth quintile, ethnicity, smoking status, arthritis, cancer, stroke, chronic lung disease, diabetes, cardiovascular diseases.

**Supplementary Figure 5. Volcano plots visualizing protein concentration associated with cross-sectional eudaimonic wellbeing, hedonic wellbeing, depressive symptoms, and life satisfaction at protein measurement from linear regressions, with multiple adjustments.**

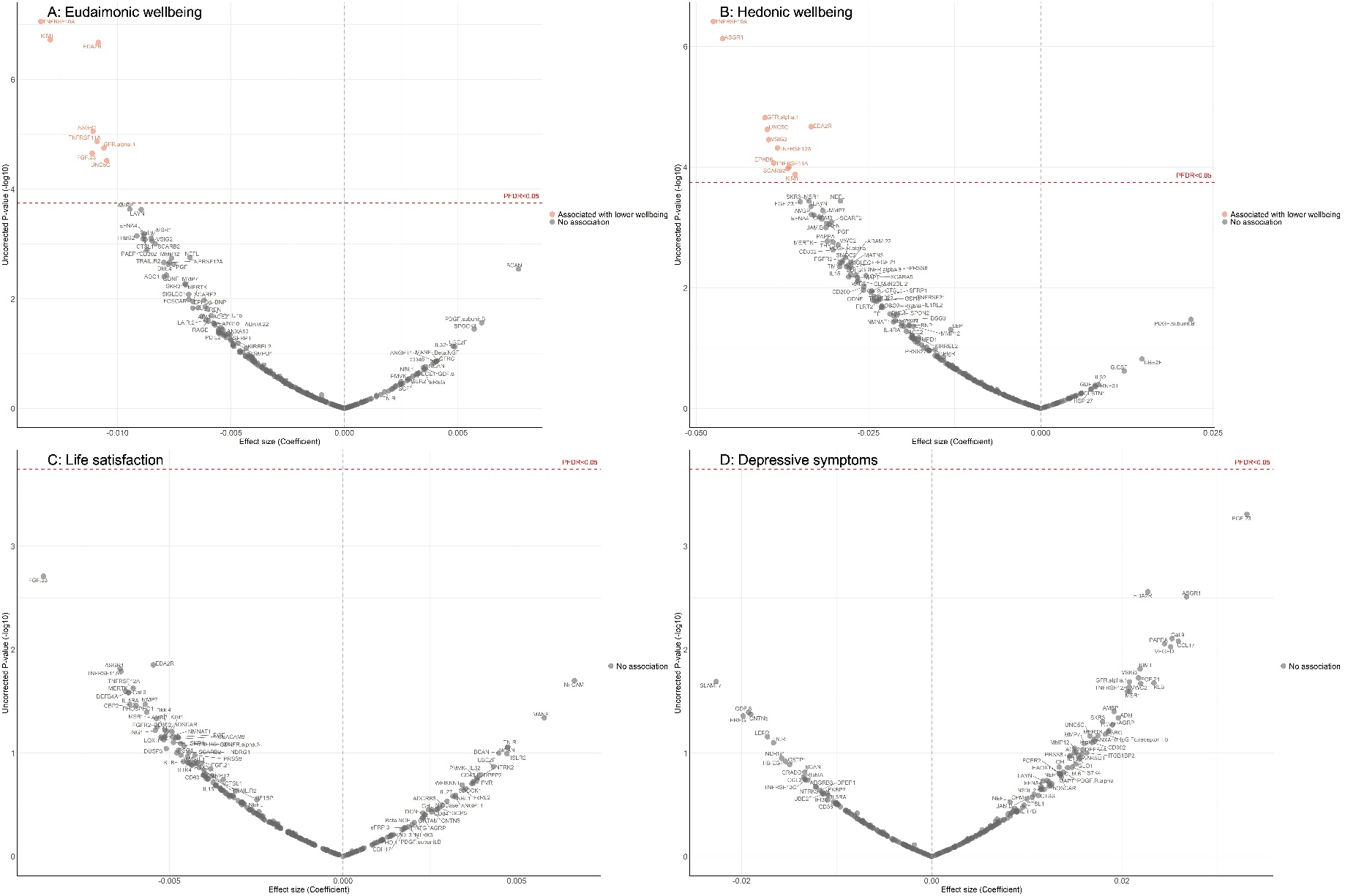

Linear regression models for protein-wellbeing associations at wave 4. Models adjusted for age, sex, wealth quintile, ethnicity, smoking status, arthritis, cancer, stroke, chronic lung disease, diabetes, cardiovascular diseases, and body mass index.

**Supplementary Figure 6. Volcano plots visualizing protein concentration associated with eudaimonic wellbeing, hedonic wellbeing, depressive symptoms, and life satisfaction after protein measurement from linear mixed effect regressions, with minimal adjustments.**
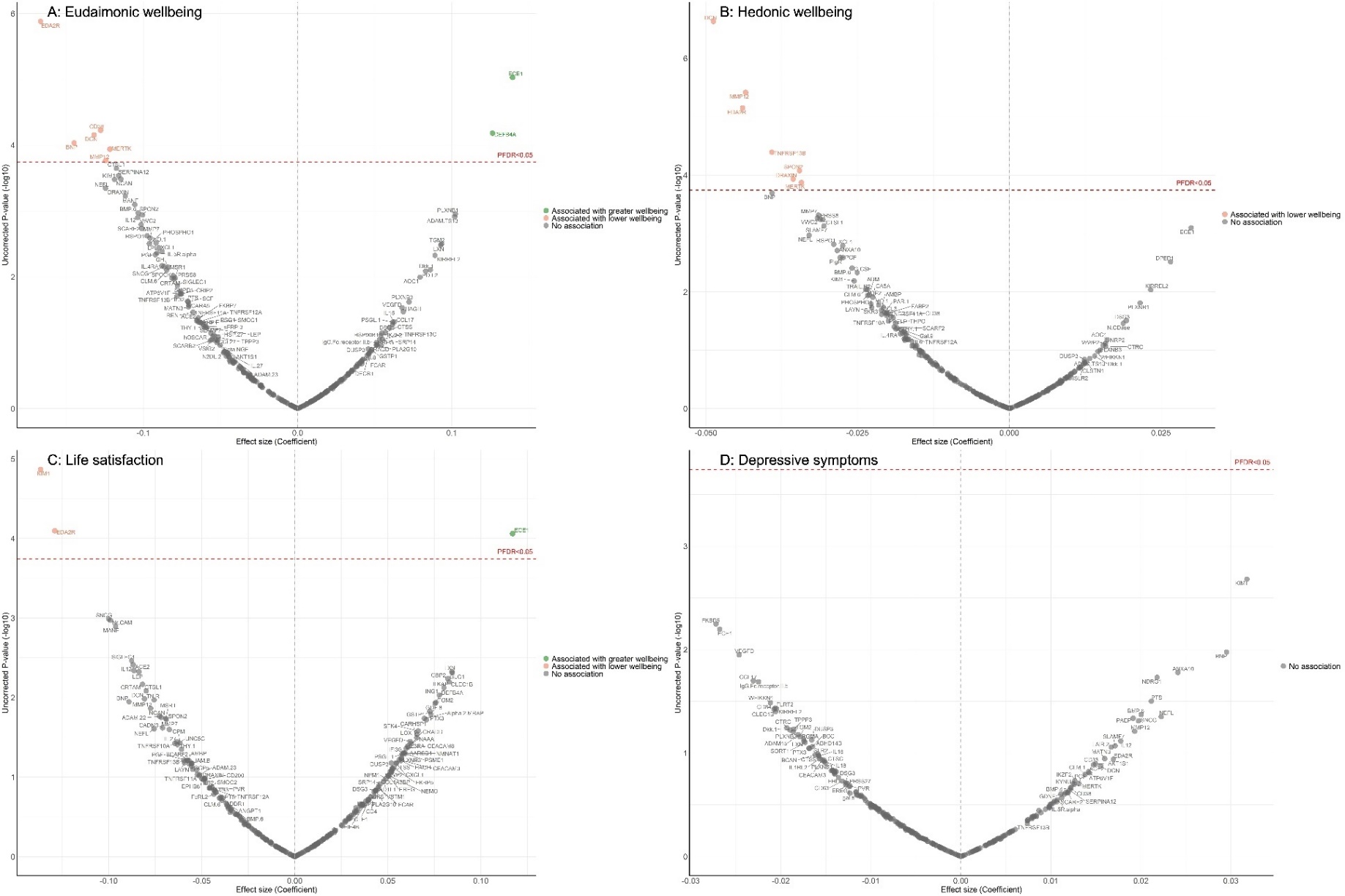

Linear mixed effect regression models for protein-wellbeing associations from wave 4 to wave 10. Models allowed for protein interacting with time since protein measurements, and adjusted for baseline age, sex, wealth quintile, ethnicity.

**Supplementary Figure 7. Volcano plots visualizing protein concentration associated with eudaimonic wellbeing, hedonic wellbeing, depressive symptoms, and life satisfaction after protein measurement from linear mixed effect regressions, with multiple adjustments.**
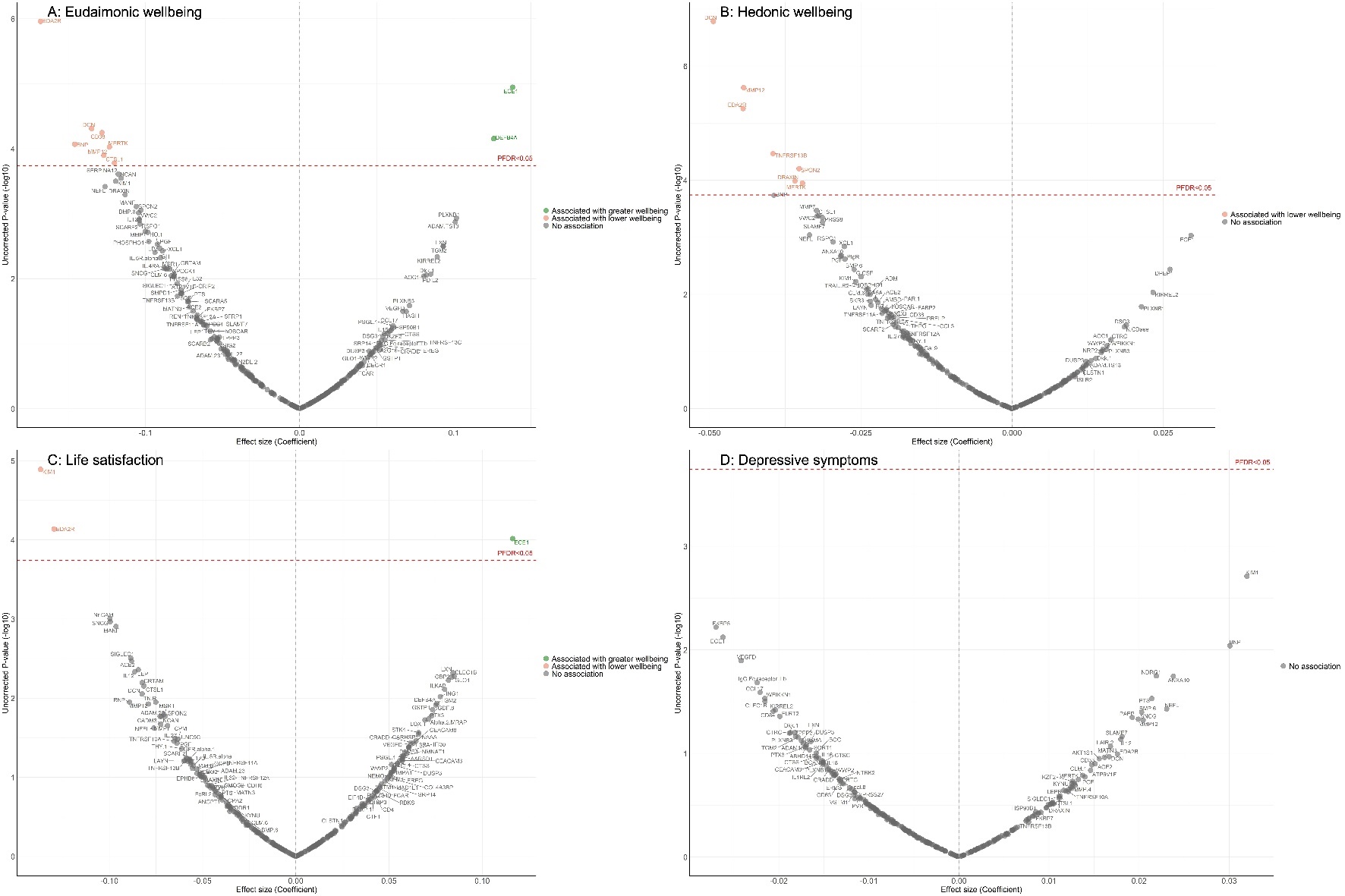

Linear mixed effect regression models for protein-wellbeing associations from wave 4 to wave 10. Models allowed for protein interacting with time since protein measurements, and adjusted for baseline age, sex, wealth quintile, ethnicity, smoking status, arthritis, cancer, stroke, chronic lung disease, diabetes, cardiovascular diseases, and body mass index.

**Supplementary Figure 8. Volcano plots visualizing protein concentration associated with eudaimonic wellbeing, hedonic wellbeing, depressive symptoms, and life satisfaction before protein measurement from linear mixed effect regressions, with minimal adjustments.**

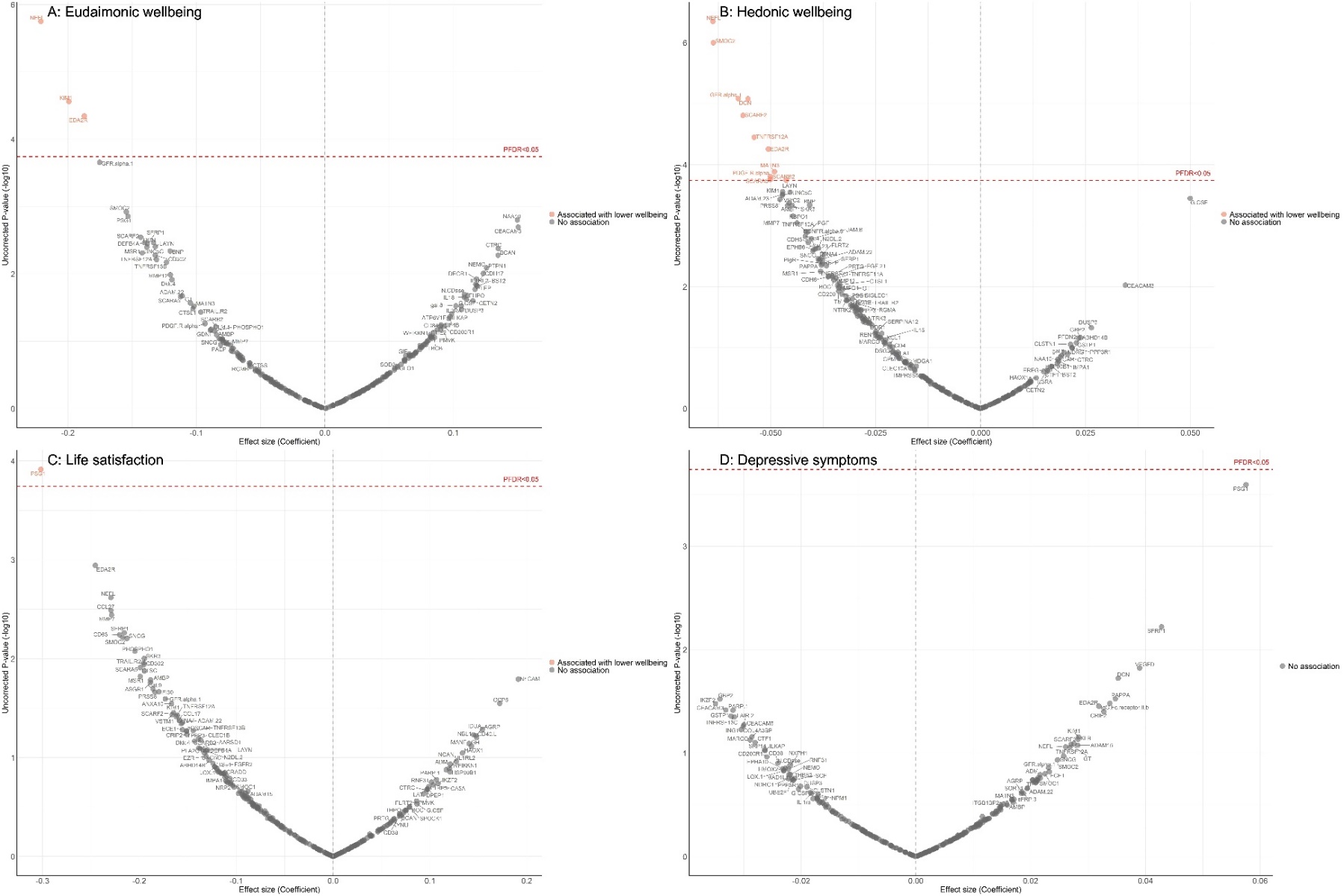

Linear mixed effect regression models for protein-wellbeing associations from wave 1 to wave 4 (except for life satisfaction, while was from wave 2 to wave 4). Models allowed for protein interacting with time since protein measurements, and adjusted for baseline age, sex, wealth quintile, ethnicity.

**Supplementary Figure 9. Volcano plots visualizing protein concentration associated with eudaimonic wellbeing, hedonic wellbeing, depressive symptoms, and life satisfaction before protein measurement from linear mixed effect regressions, with multiple adjustments.**

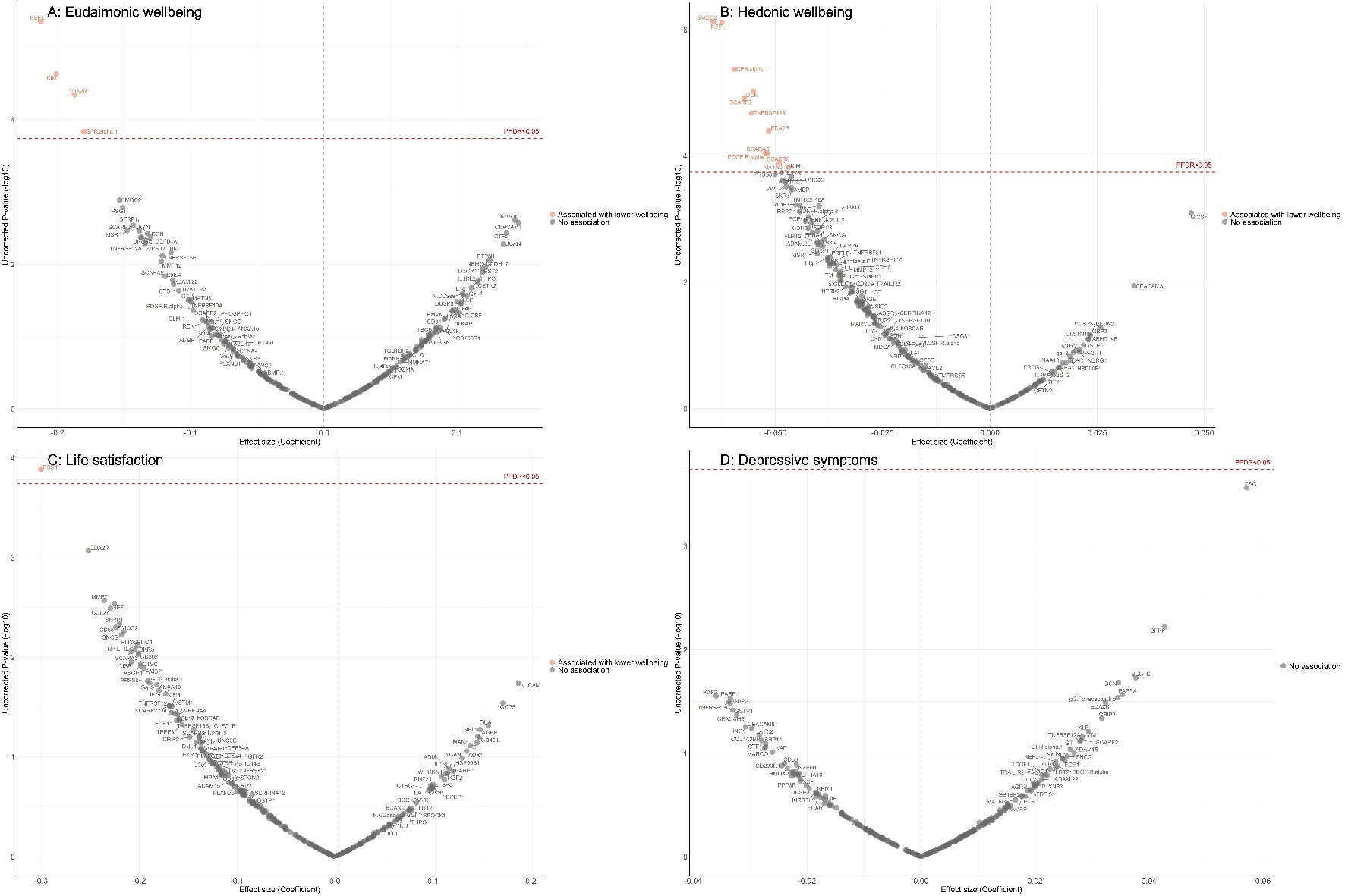

Linear mixed effect regression models for protein-wellbeing associations from wave 1 to wave 4 (except for life satisfaction, while was from wave 2 to wave 4). Models allowed for protein interacting with time since protein measurements, and adjusted for baseline age, sex, wealth quintile, ethnicity, smoking status, arthritis, cancer, stroke, chronic lung disease, diabetes, cardiovascular diseases and body mass index.
